# A plasma peptidomic signature reveals extracellular matrix remodeling and predicts prognosis in alcohol-related hepatitis

**DOI:** 10.1101/2023.12.13.23299905

**Authors:** Khaled Sayed, Christine E. Dolin, Daniel W. Wilkey, Jiang Li, Toshifumi Sato, Juliane I Beier, Josepmaria Argemi, Ramon Bataller, Abdus S Wahed, Michael L Merchant, Panayiotis V Benos, Gavin E Arteel

## Abstract

Alcohol-related hepatitis (AH) is plagued with high mortality and difficulty in identifying at-risk patients. The extracellular matrix undergoes significant remodeling during inflammatory liver injury that can be detected in biological fluids and potentially used for mortality prediction. EDTA plasma samples were collected from AH patients (n= 62); Model for End-Stage Liver Disease (MELD) score defined AH severity as moderate (12-20; n=28) and severe (>20; n=34). The peptidome data was collected by high resolution, high mass accuracy UPLC-MS. Univariate and multivariate analyses identified differentially abundant peptides, which were used for Gene Ontology, parent protein matrisomal composition and protease involvement. Machine learning methods were used on patient-specific peptidome and clinical data to develop mortality predictors. Analysis of plasma peptides from AH patients and healthy controls identified over 1,600 significant peptide features corresponding to 130 proteins. These were enriched for ECM fragments in AH samples, likely related to turnover of hepatic-derived proteins. Analysis of moderate versus severe AH peptidomes showed a shift in abundance of peptides from collagen 1A1 and fibrinogen A proteins. The dominant proteases for the AH peptidome spectrum appear to be CAPN1 and MMP12. Increase in hepatic expression of these proteases was orthogonally-validated in RNA-seq data of livers from AH patients. Causal graphical modeling identified four peptides directly linked to 90-day mortality in >90% of the learned graphs. These peptides improved the accuracy of mortality prediction over MELD score and were used to create a clinically applicable mortality prediction assay. A signature based on plasma peptidome is a novel, non-invasive method for prognosis stratification in AH patients. Our results could also lead to new mechanistic and/or surrogate biomarkers to identify new AH mechanisms.

**Lay summary:** We used degraded proteins found the blood of alcohol-related hepatitis patients to identify new potential mechanisms of injury and to predict 90 day mortality.

---

Alcohol-related hepatitis (AH) is a subacute form of alcohol-related liver disease with a high mortality rate of 30-50% at 3 months and 40% at 6 months [1, 2]. AH is characterized by hepatic decompensation, jaundice and multiple organ failure [3]. AH occurs in patients with heavy chronic alcohol consumption (80-100 g per day) and can be the first manifestation of clinically silent ALD or an exacerbation of pre-existing cirrhosis [3].

Accurately predicting AH patient outcome risks is important for clinical decision-making. For example, AH patients with higher negative outcome (e.g., mortality) risks are better candidates for corticosteroid treatment, and patients with lower risk could be candidates for long-term clinical studies [2, 3]. Currently, the best approach for predicting outcome risk is combining static scores, such as the modified Maddrey’s discriminant function (MDF), Model for End-stage Liver Disease (MELD), prognostic algorithm score constituting Age, Bilirubin, INR and Creatinine (ABIC), and/or Glasgow with the dynamic Lille scoring system [4, 5], with the MELD score favored globally [6]. These clinical scores are useful for predicting outcome risks in patients with severe AH, but are limited in predicting outcome risks in patients with moderate disease [7]. Our group demonstrated significant extracellular matrix (ECM) remodeling during inflammatory liver injury [8]. During such remodeling, altered protein turnover shifts the distribution of peptide fragments including degraded ECM in biologic fluids (e.g., plasma) [9]. Peptidomic analysis of the degraded ECM (i.e., ‘degradome’) is a useful diagnostic/prognostic tool in metastatic cancers and other diseases of ECM remodeling [9].

Probabilistic Graphical Models (PGMs), in general (i.e., “causal graphs”) have recently gained popularity, because of their simplicity and straightforward interpretability. When certain assumptions are met,[10] there are theoretical guarantees that DAGs will recover the true cause-effect associations [11, 12]. Current methods can handle mixed data types (continuous and discrete) [13, 14], and have reduced processing time [15, 16], overcoming past obstacles. Importantly, these methods can be used to build robust predictive models of an outcome [17, 18].

It was hypothesized that the severe inflammatory liver injury caused by AH would yield a unique peptidome profile in human patient plasma, and that unique ECM peptides or peptide grouping would vary between patient groups. The goals of this work were three-fold: 1) identify novel surrogate candidate biomarkers for AH, 2) develop new mechanistic hypotheses by predicting proteases that generated the observed peptidome, and 3) employ PGM to identify unique predictors of outcome from the peptidome profile (see **Figure 1** for scheme).

**Figure 1.**
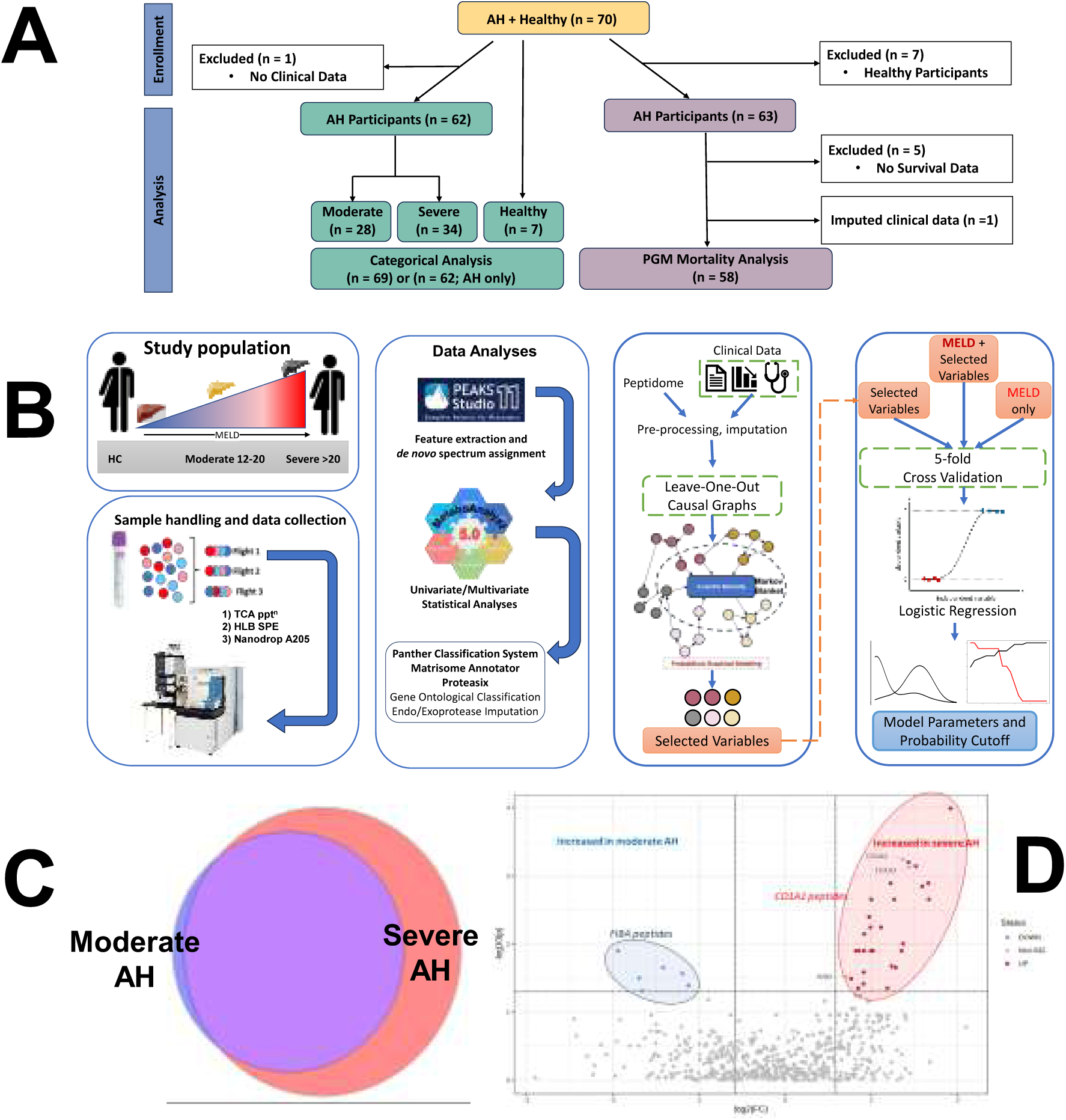
Study design and peptidome. <u>Panel A</u>: Consort Diagram. <u>Panel B</u>: Analytic workflow. Plasma proteins were precipitated with trichloroacetic acid (TCA). The peptidome was concentrated and desalted using solid phase extraction prior to data collection by high resolution, high mass accuracy UPLC-MS. Database and de novo MS spectral assignments were made using Peaks Xpro. Raw peptide abundances were normalized based on total extracted ion chromatograms (XIC) and then preprocessed within Metaboanalyst. Data were mined by univariate and multivariate statistical methods for differentially abundant peptides and peptide groups, for Gene Ontology (Panther), parent protein matrisomal composition (MatrisomeAnnotator) and for protease involvement (Proteasix). Machine learning methods were initiated with patient-specific TIC normalized peptidome and clinical scoring data (e.g., MELD, 90day mortality). Data were preprocessed to address missing values and leave-one out causal graphs to building a selected variables data set. The performance of the selected variables with or without MELD scores was compared to MELD alone using a 5-fold validation and logistic regression to establish model parameters and prediction of 90-day mortality in AH. <u>Panel C</u>: Plasma peptidome analysis by Venn diagram for prevalence (AH moderate vs. AH severe). <u>Panel D</u>: Volcano plot for significant differences (FC >±1.5; *p*<0.05). Significant peptide data points were labeled using the gene name. The analysis defines shifts of increased fibrinopeptide A (FBA) in moderate and increased collagen (e.g., CO1A1) peptides in severe AH.

## Methods

### Study participants

The University of Louisville Human Studies Committee Human approved sample collection and use of de-identified samples provided in this study. All study subjects provided informed consent prior to sample collection. All studies were conducted in compliance with the Declaration of Helsinki. A total of 70 adult male and female individuals participated in this NIH-funded study (**Figure 1A**). This investigation constitutes a single time point assessment of patients between the study subgroups. Data were collected from biobanked samples from a large national multisite clinical trial (clinicaltrials.gov: NCT01922895 and NCT01809132). Inclusion and exclusion criteria are listed in those studies. Informed consent was obtained from all study participants before collection of data and bodily samples. All AH patients were enrolled at the University of Louisville, the University of Massachusetts Medical School, the University of Texas-Southwestern and the Cleveland Clinic. AH diagnosis was done using clinical and laboratory criteria described by the NIAAA consortium on AH [19]. Individuals with liver injury met the criteria for AUD based on DSM 4 XR or DSM 5 manual. All healthy participants were recruited at the University of Louisville free of any clinically diagnosed disease (liver or organ systems) that might contribute to altered laboratory values in comparison analyses.

The subgroups included healthy participants (n=7) and AH patients (n=63; **Figure 1A**). Clinical variables did not exist for healthy controls and for one AH patient and were therefore excluded from categorical analyses (n=62). For categorical comparisons, AH patients were stratified as “moderate” (MELD=12-20; n=28) and “severe” AH (>20; n=34).[20] Out of the 63 AH patients, survival information was lacking for 5 patients, so they were excluded from the causal graphical modeling (n=58; see **Supplemental Material**). A variety of clinical data was gathered for these patients, including transaminases, alkaline phosphatase, and total bilirubin. **Table 1** shows a list of demographics and clinical data for the moderate AH, and severe AH participants.

**Table 1:**
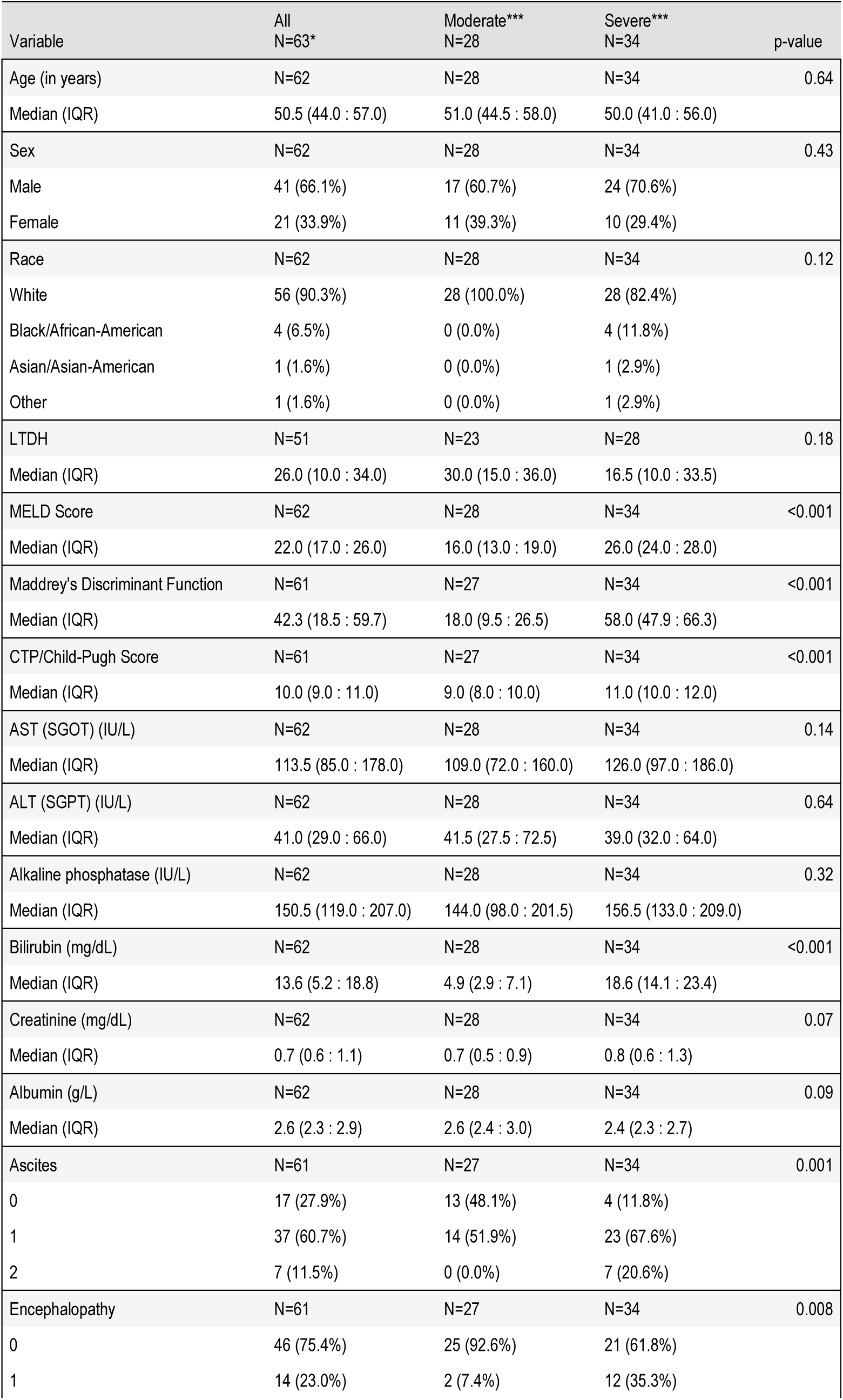
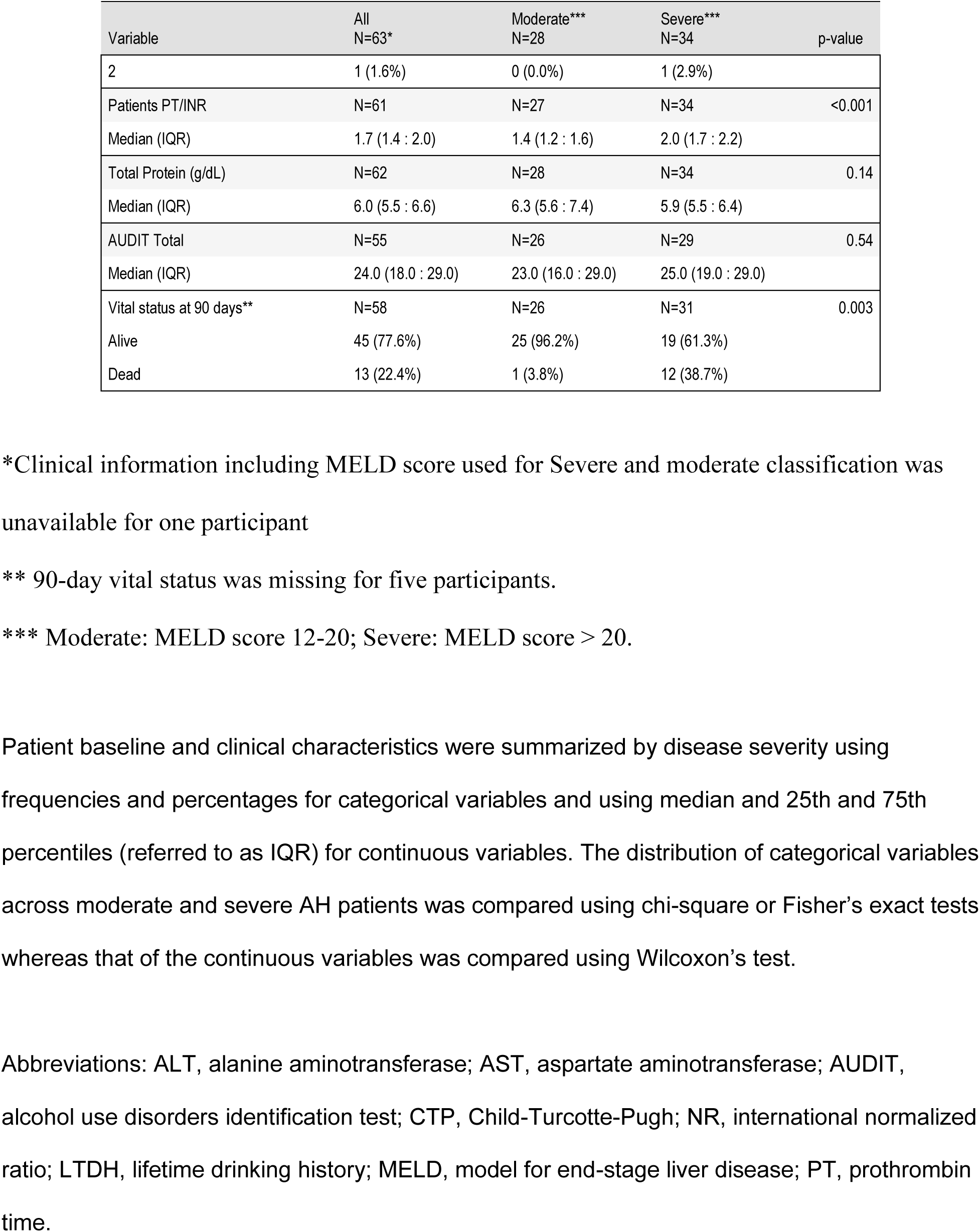
Baseline Demographic and Characteristics by Disease Severity.

### Analytical approaches, data analysis and causal graphical modeling

Peptidomic analysis of patient samples were conducted as recently described with some modifications (**Figure 1B**) [21]. That and all other detailed methods are provided in Supplemental Material.

## Results

For initial analysis, we compared the degradome in moderate (MELD 12-20) and severe (MELD >20) AH versus healthy controls. Moderate and severe AH patients did not differ significantly by age, sex, or race (**Table 1**). The median age was 51 years, 66% of patients were male, and 90% were white. In addition to MELD, severe AH patients had higher MDF (median 58 vs. 18), CTP/child-Pugh score (median 11 vs. 9), bilirubin (median 18.6 vs. 4.9 mg/dl), and INR (median 2.0 vs 1.4) and Ascites score (88.2% vs 51.9% 1-2) than the moderate AH patients (all *p*<0.001). AST and ALT did not differ significantly by MELD score severity. The peptidomic dataset consisted of 1,693 primary peptidome features identified by PEAKS X Pro, corresponding to degradation products of 134 unique proteins. There was significant qualitative overlap between peptides changed in Moderate and Severe AH (vs healthy control), as visualized by Venn Diagram (**Figure 1C**). Differences in relative peptide abundance between moderate and severe AH were determined using t-test (on preprocessed TIC-normalized data); volcano plots visualize these results (**Figure 1D**). These data demonstrate a shift toward increased relative abundances of collagen 1A1 (COL1A1) and collagen 1A2 (COL1A2) fragments and relative decreases in some fibrinogen A (FGA) peptide fragments in severe AH.

### Feature analysis of the peptidome

PCA showed that the two largest principal components account for 8.0% and 5.8% variability between the three participant categories (healthy, and moderate and severe AH; **Figure 2A**). Repeated analysis of healthy versus moderate and healthy versus severe categories demonstrated a slight increase in PC1 to explaining 10% of the data separation (**Figure 2B**). Comparison of the moderate versus severe AH samples decreased PC1 to 7% thus suggesting most of the variability in the data, could be attributed to the differences between healthy versus moderate and healthy versus severe.

**Figure 2.**
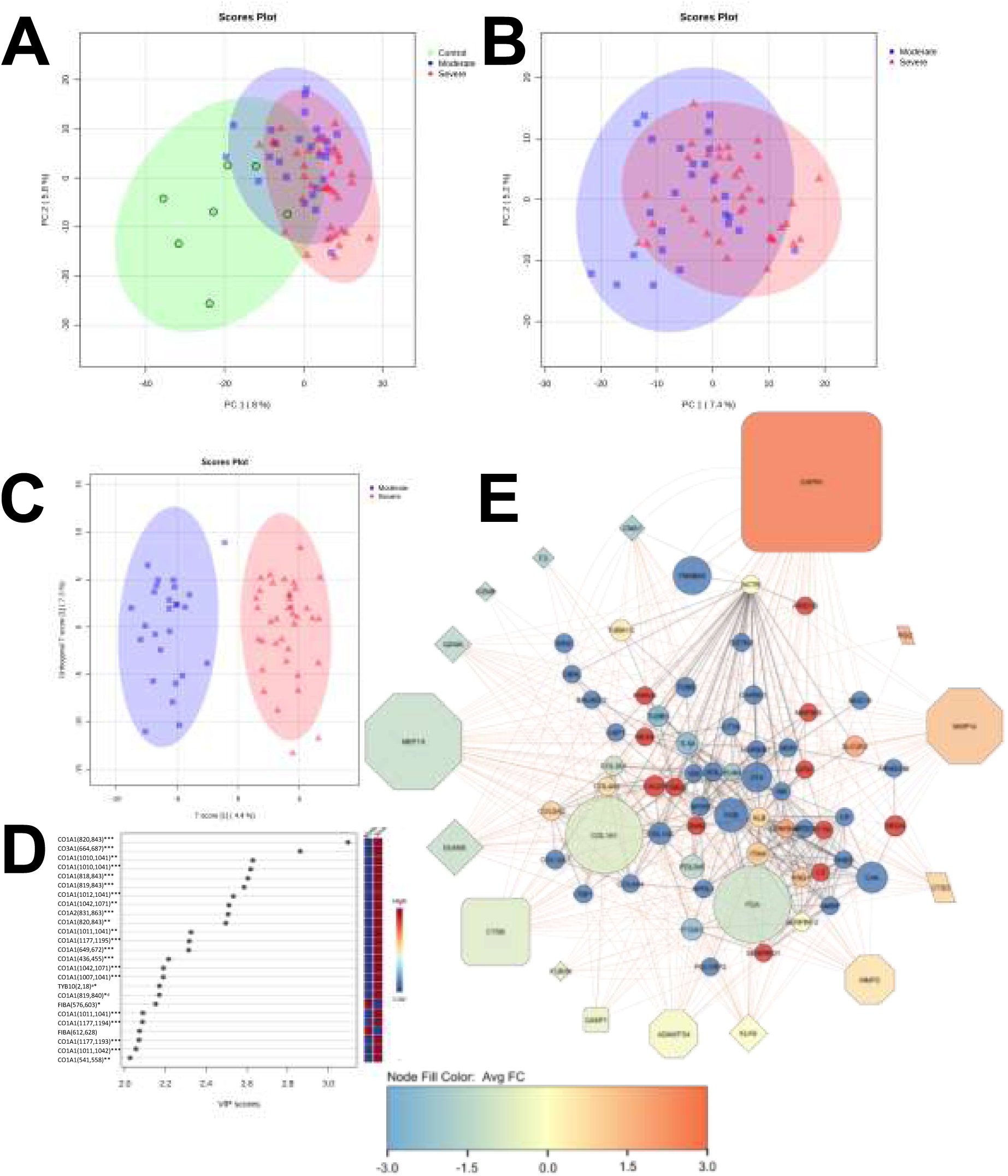
Plasma peptidome features analysis. <u>Panel A:</u> PCA analysis showing principal components PC1 and PC2 for self-sorting of healthy control (green), moderate AH (blue) and severe AH (red) samples as defined by 95% confidence intervals. Healthy control samples are resolved from AH samples. <u>Panel B</u>: Two-group analysis of moderate AH versus severe AH samples demonstrates emerging self-sorting properties of the peptidome. <u>Panel C</u>: oPLS-DA analysis comparing AH severity. Complete separation of the moderate and severe AH peptidomes is achieved using discriminate analysis. <u>Panel D</u>: Major peptide features sorted by oPLS-DA of AH samples are prolyl-hydroxylated CO1A1 fragments (severe AH) and FBA fragments (moderate). Peptide fragments defined by parent protein Gene Name, amino acid (start, stop) location, and site-specific modifications: *, prolyl hydroxylation; a, acetylation; d, dehydration <u>Panel E</u>: Cluster analysis of the peptidome/degradome in AH. The peptides significantly increased in AH were analyzed by the Proteasix (http://www.proteasix.org) algorithm using a positive predictive value (PPV) cut-off to 80%. Protein-protein interaction network analysis of regulated proteomic data sets (q-value <0.05) was performed using Search Tool for the Retrieval of Interacting Genes/Proteins, STRING v11,[23] with the highest confidence score (0.900). The resultant matrix of both Proteasix and STRING analyses were visualized using Cytoscape v3.9.1. Node sizes of the predicted proteases represented the relative frequency with which the top 16 proteases were predicted to mediate the observed cleavage (0.2-25%). Node shape for the proteases represents protease family subtype: serine (diamond), cysteine (square), aspartyl (parallelogram), and metalloproteases (octagon). Node color for protease corresponds to the Log2FC (vs healthy control) of hepatic mRNA expression from previously published work.[25] Raw data and metadata are publicly available in the Database of Genotypes and Phenotypes of the National Library of Medicine under the accession study code phs001807. Node sizes of the peptides represented the relative number of unique peptides (1-61) identified from each parent protein. Node colors of the peptides represented the median Log2FC vs healthy controls for all peptides derived from that parent protein. Solid lines depict connections between the parent proteins identified by STRING; broken lines depict predicted protease events identified by Proteasix.

#### Impact of AH on plasma peptidome profile: dominance of matrisome-derived peptides

The matrisome is an expanded definition of the ECM that also incorporates ECM-affiliated proteins and ECM-modifying proteins.[22] The gene names associated with all identified peptides (**Supplemental Table S1**) were submitted for annotation by matrisome category and division using the matrix-annotator tool *MatrisomeAnalyzer* (https://matrinet.shinyapps.io/MatrisomeAnalyzer/). The plasma peptidome comparison of the healthy versus AH samples were enriched in AH samples for peptides belonging to components of the core matrisome (collagen, ECM glycoprotein, proteoglycan) or the matrisome associated compartment (ECM regulators, ECM-affiliated proteins, secreted factors; **Supplemental Table S2**). The pattern of proteins from significantly differential abundant peptides that belong to matrisome did not differ with AH severity, comprising 60% of the total peptide signal (vs. healthy controls). The majority were defined as core matrisome proteins (collagens, ECM glycoproteins and proteoglycans) in all comparisons.

To further assess the molecular differences between the moderate and severe AH, we performed a supervised oPLS-DA score plot analysis of the plasma peptidome profiles of individual patients (**Figure 2C**) grouped by moderate and severe AH. The variation both within (7.3%) and between (4.4%) was small suggesting the differences between AH groups are driven by a small set of peptides. Variable importance plot (VIP) identified the top 25 peptides separating moderate and severe AH groups. In total, 23 of the top 25 peptides were collagen fragments. Interestingly, orthogonal partial least squared-discriminant analysis (oPLS-DA) indicated several overlapping ECM-derived peptides (e.g., degraded fibrinogen and collagen proteins) were dominant in the top-scored peptides (**Figure 2D**). These differences in plasma peptidomes suggest that biomarkers could be developed.

#### Biological pathway analyses indicate remodeling of hepatic tissue

The significantly different peptides in AH (moderate AH vs controls, severe AH vs controls) were analyzed using StringDB [23], which contains physical and functional protein-protein associations from both experimentally validated and homology-based associations. Molecular pathways related to altered metabolism and remodeling were also enriched in Gene Ontology (GO) terms for Biological Process and Cellular Component (**Supplemental Tables S3 and S4**). Alcohol-related hepatitis is a systemic disorder and extrahepatic dysfunction (e.g., skeletal muscle and kidneys) is a key driver of mortality [24]. Despite this factor, the liver was the most common solid organ enriched in the data set as determined by Brenda Tissue Ontology (BTO:0000759; **Supplemental Table S5**) in the AH-moderate and -severe versus healthy controls (*p*=5.82×10^-11^ and 4.90×10^-16^, respectively), and second only to plasma proteins (e.g., BTO:0000131; *p*= 2.22×10^-14^ and 4.39×10^-23^, respectively). Interestingly, another tissue that was highly enriched in the peptidome in AH-moderate and -severe versus healthy controls was determined to be of fetal origin (BTO:0000449; *p*=1.56×10^-9^ and *p*=7.68×10^-8^, respectively), which is in-line with previous studies (e.g., [25]).

#### Calpains and MMP-14 proteases are predicted to regulate the observed peptidome

Many proteases cleave substrates with high specificity only at certain sequence sites. Thus, information on the fragment sequence of degraded proteins can inform on proteases that may have generated this pattern. Proteasix (proteasix.org) is an open-source peptide-centric tool to predict *in silico* the proteases involved in generating these peptides [26]. **Figure 2E** shows the relative frequency (node size) with which the top 16 proteases were predicted to generate the resultant peptidome peptides by this analysis. The two top predicted upregulated proteases, Calpain −1/−2 (CAPN1/CAPN2) and MMP-14 (MMP14) were also robustly induced in publicly-available RNAseq expression data from human AH (**Figure 2E**, node color) [25]. Indeed, there was generally good concordance between the Proteasix prediction and the hepatic gene expression data from that study.

#### Peptide features that are directly linked to 90-day survival in AH

The initial analysis was to categorically describe the gestalt changes in the peptidome caused by moderate and severe AH (as determined by MELD). These results may yield useful insight for future mechanistic or interventive studies. However, as mentioned in the Introduction, a key limitation in the clinical management of AH is an accurate tool to predict outcome after clinical presentation, namely patient 90-day survival. We hypothesized that representative peptides from the peptidome could serve as surrogate biomarkers to predict this outcome.

Identifying differentially abundant peptides and pathways related to AH severity is undoubtedly useful for future studies. However, for clinical purposes, we were also interested in identifying a few peptides that could inform AH patient survival in conjunction or independently of the MELD score. To identify such potential effector peptides, we used (causal) probabilistic graphical models. Given the relatively small size of our unique dataset, we followed a leave-one-out approach, where we learned 58 graphs (one for each sample that was left out) at 10 different scarcities (see Methods). Then we counted the times, out of a total of 580 models, a particular variable was independently linked (i.e., belongs to its Markov blanket) to the 90-day mortality (binarized) variable. The graph models that did not include clinical features had four peptide fragments that consistently appeared in >90% of the graphs: X83A, X54A, X79C, and X142A. These fragments correspond to parent genes VIM, APOC1, TUBB, CALD1, respectively (**Supplemental Figure S2**). A fifth fragment (X231B, gene BIN2) appears in 76% of the models (**Supplemental Table S6**). Three of these genes (CALD1, TUBB, VIM) belong to the cytoskeleton pathway.

When we included the clinical features and the MELD score in our models, the same four fragments still appeared in >90% of the graphs, but fragment X231B appeared in 24.5% of the models (**Supplemental Table S7**). The MELD score was also found in the top informative features, as it was included in 58% of the models. We noted that MELD and X231B (BIN2) appear together in only 10 of the models (1.7%), indicating they may contain redundant information regarding the 90-day survival. **Supplemental Figure S8** shows the distribution of the identified variables within each predicted outcome group (i.e., alive vs deceased).

Additionally, we performed stepwise regression analysis on the five informative peptides to further reduce the number of relevant peptides that can be measured in a clinical setting when predicting the mortality of new patients. We found that only X54A (APOC1), X142A (CALD1), and X231B (BIN2) are the most relevant features, and these are the features we used for our predictive model.

#### Development of a three-peptide signature for clinical application

In clinical practice it is not feasible to use whole peptidome measurements as a prognostic test. Therefore, we wanted to test whether the model we learned from TIC-normalized data can be used to make predictions about survival when a limited number of peptides are measured. Thus, we simulated a clinical application of the predictive ability of the 3-peptidome signature as follows. We divided the dataset into five folds. In each fold, we used 80% of the samples (training set) to select the optimum parameters of the three peptides (“signature peptides”) through 10X cross-validation. For the training phase, we used the TIC-normalized data. The performance of the model of this fold was assessed on the raw measurements of the 20% of the left-out samples (validation set).

To avoid sample-to-sample variation in these three peptides, we used a small number of peptides to act as normalization standards. We chose the four most invariant peptides (*p*-value >0.95) with mean raw concentration >14 across all samples (“invariant peptides”) as our normalization standard. To overcome the problem of spontaneous missing values, normalization was done based on the median of the 4 invariant peptides. The selected most invariant peptides were: X38A, X157A, X61A, and X154C, corresponding to parent proteins CO3, LASP1, FETUA, and ITIH4 respectively. The median of the four invariant peptides was calculated for all samples with 3 or more non-zero values to normalize the peptides associated with mortality as shown in

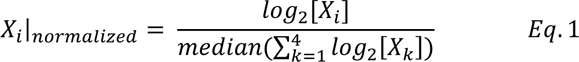

where 𝑋_𝑖_ represents a model variable and 𝑋_𝑘_ represents an invariant variable. Both 𝑋_𝑖_ and 𝑋_𝑘_ can be non-TIC or TIC-normalized peptides. It is worth mentioning that only one sample had two non-zero values. In this way, in a potential clinical application, one has to measure only these 7 peptides (3 signature and 4 invariant peptides).

Using the above procedure, we learned and compared three logistic regression models. *Model 1* consisted of the MELD score only and served as our baseline model. *Model 2* consisted of the three peptidomic features only. *Model 3* included the three peptidomic features and the MELD score. After learning the optimum weights using 10X cross-validation on the 80% of the data in each fold, we selected the optimum classification threshold as the point of intersection of sensitivity and specificity (**Figure 3A**). The threshold value where the sensitivity and specificity curves intersect was selected as the optimal threshold in each fold (**Supplemental Table S8**). The density function for the two categories (alive and deceased) across thresholds for each model is plotted in **Figure 3B**. This shows that the model only with the MELD score did not separate the two distributions well, while the peptidome models (with or without the MELD score) performed better. The validation results of each cross-validation fold are shown in **Table 2**. Like the initial training analysis (see **Supplemental Table S7**), we see that the 3-peptide model and the composite model (MELD+3-peptides) improve the MELD-only model in terms of average sensitivity, specificity, and balanced accuracy (**Figure 3C**).

**Figure 3.**
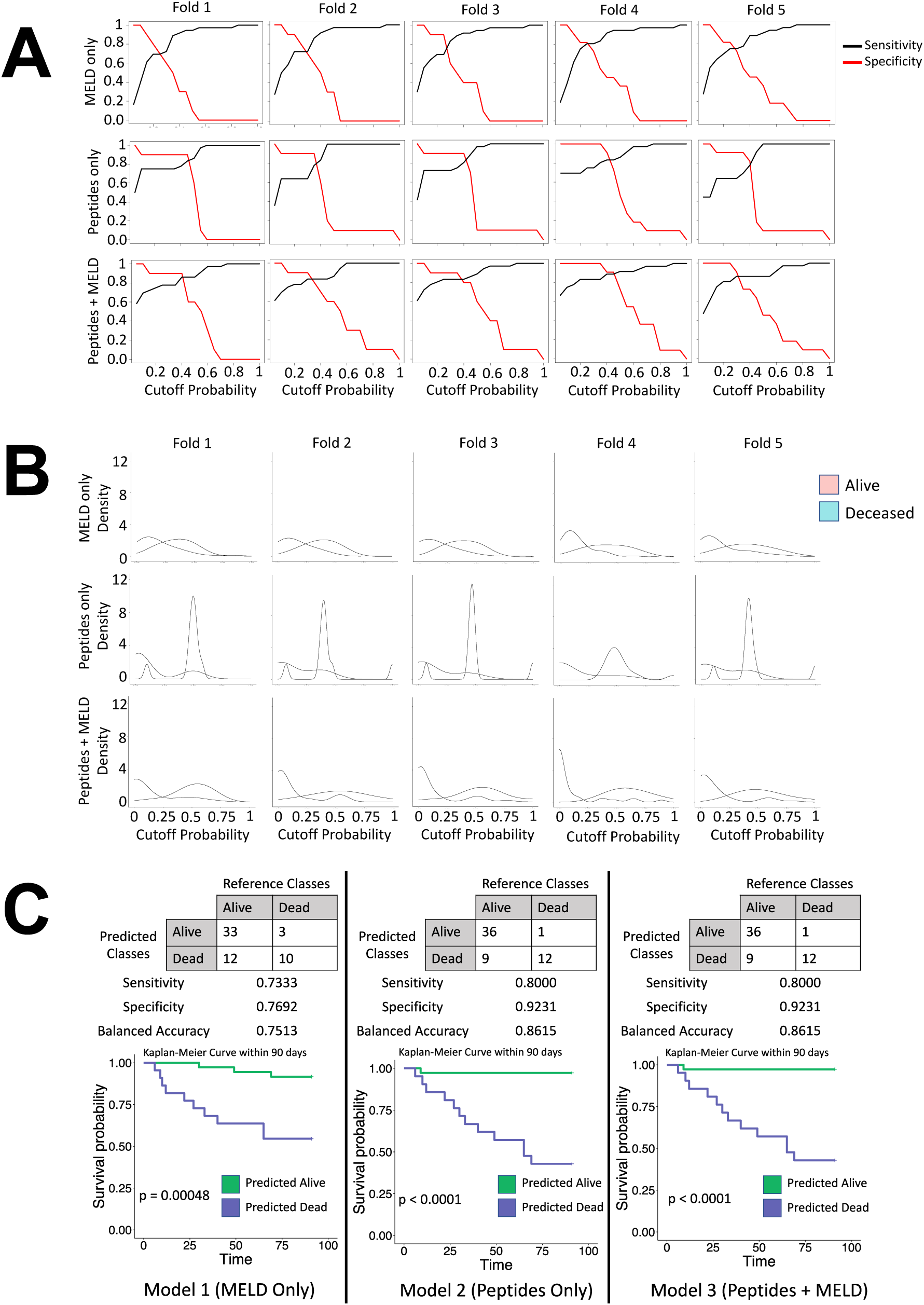
CGM modeling of the peptidome and clinical features to predict AH outcome. <u>Panel A</u>: Sensitivity and Specificity of the 5-fold cross-validation during the prediction phase of model development. X-axis: the threshold used in the parameter sweep (range 0.1-1.0). The intersection of sensitivity and specificity was used to determine the optimal threshold for each fold in each model. <u>Panel B</u>: Density distribution of 90-day survival classification over different cutoff probability thresholds. Model 2 and Model 3 offer better separation of the two categories than the MELD score alone. <u>Panel C</u>: Comparison of model performance using the complete dataset. The tables show the number of correctly and incorrectly classified samples. Sensitivity, specificity, and balanced accuracy summarize these results. Kaplan-Maier survival plots depict the discrimination ability of the three models.

**Table 2:** Causal graphical modeling validation results. Sensitivity, specificity, and accuracy values are provided for each fold of each model, while average values are also given. Red/bold font designates the maximum average value of each metric.

|  |  | Fold 1 | Fold 2 | Fold 3 | Fold 4 | Fold 5 | Average |
| --- | --- | --- | --- | --- | --- | --- | --- |
| Model 1<br>(MELD only) | Sensitivity | 0.8889 | 0.8889 | 0.8889 | 0.4444 | 0.6667 | 0.7556 |
|  | Specificity | 1.0000 | 0.3333 | 0.3333 | 1.0000 | 1.0000 | 0.7333 |
|  | Accuracy | 0.9444 | 0.6111 | 0.6111 | 0.7222 | 0.8333 | 0.7444 |
| Model 2<br>(Peptides only) | Sensitivity | 0.7778 | 0.8889 | 0.6667 | 0.6667 | 1.0000 | 0.8000 |
|  | Specificity | 1.0000 | 0.6667 | 0.6667 | 0.5000 | 1.0000 | **0.7666** |
|  | Accuracy | 0.8889 | 0.7778 | 0.6667 | 0.5833 | 1.0000 | 0.7833 |
| Model 3<br>(Peptides + MELD) | Sensitivity | 0.8889 | 0.8889 | 0.8889 | 0.7778 | 0.7778 | **0.8444** |
|  | Specificity | 1.000 | 1.0000 | 0.3333 | 0.5000 | 1.0000 | **0.7666** |
|  | Accuracy | 0.9444 | 0.9444 | 0.6111 | 0.6389 | 0.8889 | **0.8055** |

For the final model to be used in future clinical settings, we compared the three models, trained on the full dataset (parameter setting through 10× cross-validation on the TIC-normalized data), and tested them using the raw data (normalized by the median of the invariant peptides). Confusion matrices, sensitivity, specificity, balanced accuracy, and survival curves, obtained for each model when the models were tested using non-TIC normalized data, are presented in **Figure 3C**. Consistently with the results above, the highest performance measures were obtained for Model 3 whereas the lowest performance measures were obtained for the MELD-only model (Model 1). Additionally, Models 2 and 3 split the data into two clusters (i.e., Predicted Alive and Predicted Deceased) with 12 out of 13 Deceased samples grouped in the Predicted Deceased cluster. Model 3 outperformed Model 2 in detecting the Alive samples with a sensitivity of 80%. Finally, the survival curves in **Figure 3C** show that Models 2 and 3 can predict the survival of AH patients better than Model 1 which is based on the MELD score only. The distribution of the final model variables (i.e., X54A, X142A, and X231B) over the predicted classes obtained by each model is presented in **Supplemental Figure S3**. Additionally, the demographics and clinical characteristics of each predicted class in each model are shown in **Supplemental Tables S9-S11.**

## Discussion

Both the AH diagnosis and prognosis could be impacted by the development of more sensitive and specific surrogate candidate biomarkers. Our group previously demonstrated that inflammatory stress causes the hepatic ECM to undergo dynamic transitional remodeling [8]. Others have shown that ECM remodeling causes degradation products to be secreted into the blood and that analysis is a useful prognostic tool in diseases [9]. Therefore, here we aim to study the plasma ECM peptidome changes with AH severity and to develop a new, clinically useful method, to predict 90-day survival in AH patients.

Informatics analysis of these peptides demonstrated an enrichment of ECM fragments in the AH patient samples that is likely related to the turnover of hepatic proteins. Six fibrinopeptide A peptides were more abundant in plasma from moderate AH patients (compared to severe AH), whereas 27 peptides (24 COL1A1 fragments, one COL1A2, one COL1A3, and one BIN2 peptide fragment) were more abundant in plasma from severe AH patients. The findings of increased collagen in more severe AH is in line with previous studies indicating that underlying fibrosis drives AH prognosis [27]. Using a discriminant analysis (oPLS-DA; **Figure 2C**) the moderate and severe AH samples were well resolved into two sample groups and the VIP scores with a similar pattern (**Figure 2D**).

Analysis of this spectrum also identified 2 proteases that appear to be dominant in generating the pattern associated with AH (CAPN1 and MMP12); the increase in hepatic expression of these proteases was orthogonally validated in a separate analysis of publicly available bulk RNA-seq data of livers from AH patients (**Figure 2E**). Little is known regarding the role of MMP14 in liver disease. Recent work by this group showed that CAPN2 is progressively induced in post-transplant NASH fibrosis severity [28]. These proteases have not been identified to be involved in AH prior to this work. Future studies should investigate this finding further.

Besides investigating the molecular mechanisms of AH progression, we also wanted to develop a predictor for 90-day survival (outcome) using PGM. Significantly, the variables selected through this process provide independent information about 90-day survival, and collectively are the most informative variable set, even though some of them, individually, may not appear to be significantly different in the two categories (alive/deceased; **Supplemental Figure S2**). The final models showed that the 3-peptide model was equally accurate in predicting 90-day mortality as the composite one (MELD+3-peptides; **Figure 3C**). One peptide (APOC1) has been shown to be a marker of AH severity in a recent plasma proteomic study [29]. The other two peptides (CALD1 and BIN2) are both cytoskeletal proteins that have not been previously associated with AH and warrant further investigation.

While our goal was to differentiate moderate and severe AH and AH outcome (i.e., mortality), we acknowledge the ratio of HC to AH in our cohort was imbalanced in “n” values and this may be insufficient to adequately power the study. Additionally, the potential for a “clinical site effect” may be present, although the AH samples were collected from multiple institutions. Despite these study limitations, the statistical modeling and informatics filtering of the peptidomics data supports the hypothesis that the ECM plasma peptidome is associated with the AH spectrum. These patterns of select peptidome “features” can be investigated further in future studies as biomarkers for AH severity and outcome.

Another important consideration for surrogate biomarker discovery is the impact of the sample preparation method on the peptidome. Here, we used K3EDTA plasma, which inhibits coagulation-driven proteases that are dependent on divalent cations. K3EDTA is an anticoagulant of choice for plasma proteomic studies, in general, and peptidomics in particular [30]. Although direct comparisons between plasma preparations were not performed, several of the peptides identified as key biomarkers for AH outcome in this study (e.g., vimentin) have an affinity for heparin [31], and thus may not be present in studies involving heparinized plasma. Therefore, these study results are applicable at this point with use in studies of EDTA-based plasma samples.

Taken together, the results of this study indicate that analysis of the plasma peptidome can yield useful new information on mechanism and outcome prediction in AH. This study validates previous mechanistic findings (e.g., ECM remodeling and fetal-like reprogramming), as well as identifies new potential “players” at the level of degraded proteins and proteases that may generate these signals. Moreover, our curated algorithm identified by CGM proved to be superior to MELD both in sensitivity and specificity to predict mortality in AH. Future studies will investigate these prospects further.

## Acknowledgements

Access to healthy control and AH consortium samples provided by Craig McClain, MD and Vatsalya Vatsalya, MD through the University of Louisville Alcohol Research Center (P50 AA024337). Supported, in part, by grants from NIH (R01 DK130294, R01 AA021978, R01 HL157879, R01 HL127349, P20 GM113226, P30 DK120531).

## Author Contributions

KS: Visualization, Investigation, Validation, Formal Analysis, Writing-Original Draft. CED: Investigation, Validation, Formal Analysis, Visualization, Writing-Original Draft. DWW: Investigation, Validation. JL: Visualization, Investigation, Validation, Formal Analysis. TS: Investigation, Validation, Formal Analysis. JIB: Investigation, Validation, Formal Analysis. JA: Visualization, Investigation, Resources. RB: Visualization, Investigation, Resources. ASW: Visualization, Formal Analysis, Writing-Review and Editing.. MLM: Project Administration, Visualization, Conceptualization, Investigation, Supervision, Writing-Review and Editing, Funding acquisition, Resources. PVB: Project Administration, Visualization, Conceptualization, Investigation, Supervision, Writing-Review and Editing, Funding acquisition, Resources. GEA: Project Administration, Visualization, Conceptualization, Investigation, Supervision, Writing-Review and Editing, Funding acquisition, Resources.

## Data availability statement

Proteomic files were deposited in MassIVE (http://massive.ucsd.edu/) as study (MassIVE MSV000093513) entitled “Alcoholic hepatitis plasma degradome”. Data include (A) the primary data files (.RAW), (B) peak list files (.mzML), (C) sample key, (D) the sequence databases (human UniprotKB reviewed reference proteomes), and (E) excel files containing Peaksdb results for de novo peptide sequence assignment. The shared data will be released from private embargo for public access upon the manuscript’s acceptance for publication. All other data will be made available on request.

## Additional Information

The authors declare that they have no known competing financial interests or personal relationships that could have appeared to influence the work reported in this paper.

## Abbreviations

ABIC: age, bilirubin, INR and creatinine
AH: alcohol-related hepatitis
ALD: alcohol-related liver disease
ALT: alanine aminotransferase
AP: alkaline phosphatase
ASH: alcohol-related steatohepatitis
AST: aspartate aminotransferase
AUD: alcohol use disorder
AUDIT: alcohol use disorders identification test
BMI: body mass index
BTO: Brenda tissue ontology
CTP: Child-Turcotte-Pugh
DAG: directed acyclic graphs
DF: discriminant function
DSM: diagnostic and statistical manual of mental disorders
ECM: extracellular matrix
FGES: fast greedy equivalence search
FLIGHT: functional liver-image guided therapy
GO: gene ontology
LC-MS/MS: liquid Chromatography with tandem mass spectrometry
LTDH: lifetime drinking history
MELD: model for end-stage liver disease
PCA: principal component analysis
PGM: probabilistic graphical models
oPLS-DA: orthogonal partial least squared-discriminant analysis
TIC: total ion chromatogram
VIP: variable importance plot.

