## Supplemental Materials for "A plasma peptidomic signature reveals extracellular matrix remodeling and predicts prognosis in alcohol-related hepatitis"

[1]Department of Epidemiology, University of Florida, Gainesville, Florida, USA.
[2]Department of Electrical & Computer Engineering and Computer Science, University of New Haven, West Haven, Connecticut, USA.
[3]Department of Medicine, University of Louisville, Louisville, Kentucky, USA.
[4]Department of Medicine, Division of Gastroenterology, Hepatology and Nutrition, University of Pittsburgh, Pittsburgh, PA, USA.
[5]Pittsburgh Liver Research Center, University of Pittsburgh, Pittsburgh, PA, USA.
[6]Department of Internal Medicine, Clinical University of Navarra, Navarra, Spain.
[7]Liver Unit, Hospital Clinic. Institut d'Investigacions Biomèdiques August Pi i Sunyer (IDIBAPS), Barcelona, Spain.
[8]Department of Biostatistics and Computational Biology, University of Rochester, Rochester, NY, USA.

#equally contributing first authors

*equally contributing senior authors

Running title: The blood peptidome of alcohol-related hepatitis.

Keywords: causal models, protein degradomics, extracellular matrix, ALD, LC-MS/MS

Send all correspondence to: Gavin E. Arteel, PhD, FAASLD
Thomas E. Starzl Biomedical Science Tower
West 1144
200 Lothrop Street
Pittsburgh, PA 15213


**Abbreviations**: ABIC, age, bilirubin, INR and creatinine; AH, alcohol-related hepatitis; ALD, alcohol-related liver disease; ALT, alanine aminotransferase; AP, alkaline phosphatase; ASH, alcohol-related steatohepatitis; AST, aspartate aminotransferase; AUD, alcohol use disorder; AUDIT, alcohol use disorders identification test; BMI, body mass index; BTO, Brenda tissue ontology; CTP, Child-Turcotte-Pugh; DAG, directed acyclic graphs; DF, discriminant function; DSM, diagnostic and statistical manual of mental disorders; ECM, extracellular matrix; FGES, fast greedy equivalence search; FLIGHT, functional liver-image guided therapy; GO, gene ontology; LC-MS/MS, liquid Chromatography with tandem mass spectrometry; LTDH, lifetime drinking history; MELD, model for end-stage liver disease; PCA, principal component analysis; PGM, probabilistic graphical models; oPLS-DA, orthogonal partial least squared-discriminant analysis; TIC, total ion chromatogram; VIP, variable importance plot.

**Supplemental Detailed Methods**

Clinical variables. Clinical data included participant demographics [age, sex, body mass index (BMI)], medical assessments at admission (with consent) to exclude any condition that might affect liver tests and medical history. Confirmatory tests for AH (laboratory and imaging), and markers of liver disease severity [Child-Turcotte-Pugh (CTP), MELD, Maddrey DF] were also obtained. Lab tests [Creatinine, Albumin, Aspartate Transferase (AST), alanine aminotransferase (ALT), alkaline phosphatase (AP), Encephalopathy, Ascites, Total Protein] and *Functional liver-image guided hepatic therapy* (FLIGHT) were also included. Measures of severity of ALD, and drinking history [using the Alcohol Use Disorders Identification Test (AUDIT [1]) and lifetime drinking history (LTDH ][2]) were collected in all AH patients. Patient clinical endpoints: Forty-five patients in the cohort lived more than 90 days (labeled as Alive) and 13 patients died (labeled as Deceased) within the first 90 days of blood sampling.

Peptide extraction from plasma. Bioreposited patient plasma samples (100 µL) were thawed and aliquoted clean 1.5mL microtube and diluted with an equal volume of K3-EDTA added from fresh BD Vacutainer® tubes. Freeze-thaw cycles were avoided to maintain the integrity of the plasma. Intact plasma proteins were precipitated by adding 200 µL of ice-cold 20% w/v trichloroacetic acid in LC-MS/MS grade water for 5min followed by vortexing and incubation at 4°C for 60min. Precipitated plasma proteins were pelleted by two centrifugation steps at 16,000xg, 4°C for 10min and the peptidome-containing supernatant was transferred to a clean tube after each clarification step. Peptides were extracted from the supernatant using solid phase extraction (Oasis HLB µElution 30µm/2mg plate, Waters, Milford, MA, USA) using a Presston 100 Positive Pressure Manifold (Phenomenex, Torrance, CA, USA) using the manufacturer's protocol. The µElution plate was sequentially rinsed (200 µL) three times at a flow rate of 1mL/min with the loading buffer (5% v/v acetonitrile / 0.1% v/v TFA) and elution buffers (40% v/v can / 0.1% v/v TFA). The sample was loaded, rinsed five times (200μL) with loading buffer, and eluted three times (50μL) with elution buffer. The eluates were combined, dried in a SpeedVac, and stored at -80°C. For LC-MS analysis, the dried HLB eluates were dissolved in 25μL 2% v/v acetonitrile / 0.1% v/v formic acid and the peptide concentration was estimated with a NanoDrop 2000 (ThermoFisher Scientific, Waltham, MA, USA) using absorbance at 205 nm. Samples were diluted to 0.1μg/μL (by A205 absorbance) and 0.5x iRT retention time normalization standards (Biognosys, Schlieren, Switzerland). 2μL of each sample was analyzed by LC-MS/MS.

UPLC-MS Analysis. Samples were loaded onto Accclaim PepMap 100 75μm x 2cm, nanoViper (C18, 3μm, 100Å) trap then onto an Acclaim PepMap RSLC 75μm x 50cm, nanoViper (C18, 2μm, 100Å) separating column (ThermoFisher) heated at 50°C using an EASY-nLC 1000 UHPLC system (ThermoFisher) with a binary gradient (solvent A, 2% v/v acetonitrile / 0.1% v/v formic acid; solvent B, 80% v/v acetonitrile / 0.1% v/v formic acid). Peptidomic samples were separated with a 60min linear gradient from 0% to 55% B at 250nL/min, followed by a 5min linear gradient from 55% to 95% B with a flow ramp from 250 to 300nL/min, and then with a 10min wash with 95% B at 300nL/min. Elutes were introduced into the MS with a 40mm stainless steel emitter (ThermoFisher) positioned by a Nanospray Flex source (ThermoFisher) near the ion transfer capillary of the mass spectrometer. The ion transfer capillary temperature was set at 225°C, and the spray voltage at 1.75kV.

An Orbitrap Elite – ETD mass spectrometer (ThermoFisher) was used to collect data from the LC eluate. An Nth Order Double Play was created in Xcalibur v2.2 (ThermoFisher). Scan event one obtained an FTMS MS1 scan (normal mass range; 240,000 resolution, full scan type, positive polarity, profile data type) for the range 300-2000m/z. Scan event two obtained ITMS MS2 scans (normal mass range, rapid scan rate, centroid data type) on up to twenty peaks that had a minimum signal threshold of 5,000 counts from scan event one. The lock mass option was enabled (0% lock mass abundance) using the 371.101236m/z polysiloxane peak as an internal calibrant.

Data preprocessing. Identified peptides were quantified by PEAKS using extracted ion chromatogram area and then normalized by total ion chromatogram (TIC) area. To address the issue of peptide degradation while circulating in the blood, the peptidome data was grouped, based on overlapping peptide sequences (see “Sequences” in **Supplemental Table S1**). Area data for these groups were summed and the grouped peptidomic features were $\log_2$ transformed (+1 pseudocount).

Peptidomic Data Analysis. RAW data files were searched in Peaks Studio 8.5 (Bioinformatics Solutions Inc., Waterloo, ON, Canada) using the Denovo, PeaksDB, PeaksPTM, and Label Free Q algorithms and a protein database containing UniprotKB human-reviewed canonical and isoform sequences downloaded on 5/29/2018 plus iRT peptide sequences. No enzyme was specified for the search and Oxidation(M) was set as a variable modification. Fragment tolerance was 0.5Da and parent tolerance was 15ppm (monoisotopic). For the PeaksPTM algorithm, only the following Peaks 8.5 common PTMs were considered: Acetylation (K), Acetylation (Protein N-term), Amidation, Carbamidomethylation, Carbamylation, Carboxymethyl, Citrullination, Deamidation (NQ), Homoserine, Homoserine lactone, Phosphorylation (STY), Dehydration, Pyro-glu from E, Pyro-glu from Q, Sodium adduct, Oxidation (HW), Oxidation (M), Oxidation or Hydroxylation, Dimethylation(KR), Betamethylthiolation, Hexose (NSY), HexNAcylation (N), FAD, Guanidination, 4-hydroxynonenal (HNE), Formylation, Formylation (Protein N-term), Carboxylation (E), Ammonia-loss (C@N-term), Ammonia-loss (Protein N-term), and Dioxidation (M). FDR estimation was enabled in the PeaksDB and PeaksPTM algorithms. The Label Free Q algorithm was used with a mass error tolerance of 10ppm, and a retention time shift tolerance of 6.0min and an FDR threshold of 1%. A peptide.csv file was exported from the Label-Free Q result for curation in Microsoft Excel. Normalization was done using TIC.

High-confidence peptide assignments (Peaks Studio X criteria –logP scores with FDR threshold 1%) were exported into comma-separated values files (.csv) for upload and analysis by the Proteasix (http://www.proteasix.org) algorithm using the 'Peptide-centric' prediction tool based on curated, known and observed cleavage events enabling assignment of protease identities from the MEROPS database as described previously [3]. The positive predictive value (PPV) cut-off for the prediction algorithm output was set to 80%. Protein-protein interaction network analysis of regulated proteomic data sets (q-value <0.05) was performed using Search Tool for the Retrieval of Interacting Genes/Proteins, STRING v11 [4], with the highest confidence score (0.900). The resultant matrix of both Proteasix and STRING analyses were visualized using Cytoscape v3.9.1. Node sizes of the peptides represented the relative number of unique peptides (1-56) identified from each parent protein. Node color of the peptides represented the median Log2FC (AH vs Healthy controls) derived from that parent protein. Node sizes of the predicted proteases represented the relative frequency with which the top 16 proteases were predicted to mediate the observed cleavage (0.2-27%). Node color of the proteases represented the Log2FC (severe AH vs Healthy controls) of hepatic mRNA expression derived from secondary analysis of RNA sequencing of published data (see below).

Differentially abundant peptide analysis. Differentially abundant peptide analyses were conducted using normalized individual peptide TIC areas using the toolsets available on the website Metaboanalyst (https://www.metaboanalyst.ca/). To address the non-normal distribution of peptide abundances the peptide data were normalized within Metaboanalyst using the following approach (**Supplemental Figure S1**). Peptides with constant or single values as well as features with more than 50% missing values across all groups were removed. The remaining missing values were imputed using singular value decomposition. Data were filtered by interquartile ranges, quantile normalized, log10 transformed and pareto scaling. Data were analyzed using ANOVA to estimate differences between healthy controls, moderate and severe AH patient peptidomes using an un-adjusted p-value of 0.05 to define significance. Post-hoc t-test and volcano plots using adjusted p-values (q-value=0.05) and Log2FC (1.5) to define significant differentially abundant peptide features. Principal Component Analysis (PCA) and orthogonal Partial Least Squared-Discriminant Analysis (oPLS-DA) were used to estimate the patient peptidomes ability to self-sort the patient samples in an unbiased and discriminant fashion. The oPLS-DA variable importance plot (VIP) plots (based on ANOVA scores for the top 25 features and VIP scores > 1.0) were used to determine peptide features responsible for the greatest separation of patient peptidome groups.

Publicly available Liver RNA Sequencing Analysis. RNA sequencing data were obtained from normal livers (n = 10) and from biopsies of patients with early silent ALD (n = 12), nonsevere AH (n = 9), and severe AH (n = 28); the processing, sequencing, and clinical data have been described previously [5], and the raw data and metadata are publicly available in the Database of Genotypes and Phenotypes of the National Library of Medicine under the accession study code phs001807. v1. p1 (https://www.ncbi.nlm.nih.gov/projects/gap/cgi-bin/study.cgi?study_id=phs001807.v1.p1, last accessed June 6, 2023).

Predictive model data analysis. Causal graphs are versatile machine learning methods that can be used to identify potential cause-effect relationships among the dataset variables,[6, 7] generate hypotheses,[8] and select variables for efficient prediction of an outcome.[9] We used the Fast Greedy Equivalence Search (FGES) algorithm to generate the probabilistic graphs at penalty discount (sparsity) values from 1-10. The graphs were used to select informative peptides for AH-related mortality. Given that colinear features are problematic for graphical models, we grouped features with correlation of ≥0.3, p-value<0.05 (function "correlate", R package "corrr"), and randomly selected a representative of each group to be used in the model. The peptides were then combined with clinical data and missing clinical values (n=1) were imputed (function "knn.impute", R package "bnstruct", k = 5). Invariant peptides (to be used for calibration of the clinical risk model) were selected using the limma::lmFit function (*p*-value >0.95 and mean concentration >14). **Figure 1** highlights the overall procedure we followed to develop a model for predicting the 90-day survival of AH patients.

**Supplemental Results.**

**Table S1** lists all peptides significantly changed in AH-moderate and -severe (versus healthy control), as described in Experimental Procedures. **Table S2** summarizes the distribution of non-ECM and 'matrisome' peptides significantly changed in AH -moderate and severe (versus healthy control). **Tables S3 and S4** list the Gene Ontology (GO) terms Biological Process (GO:0008150; **Table S3**) and for Cellular Component (GO:0005575; **Table S4**) for significantly changed peptides in AH-moderate and -severe versus healthy controls. Table S5 lists the tissue enrichment by Brenda Tissue Ontology (BTO) for significantly changed peptides in AH-moderate and -severe versus healthy controls. **Table S6 and S7** summarize the frequency of variable appearances in the Markov blanket of the 90-day mortality using Leave-One-Out cross-validation on the dataset with peptidome features only (S5) and peptidome features and clinical variables (S6), as described in Experimental Procedures. **Tables S8-S10** summarize the demographics and clinical characteristics of the predicted class by models 1-3, respectively, as summarized in the Methods.

**Figure S1** shows the result of TIC normalization of the peptide results. **Figure S2** shows the separation of MELD and initial candidate peptides during the training phase of alive and dead AH patients. **Figure S3** shows the distribution of MELD and final 3 peptides using causal modeling based on MELD only (Model 1), peptidomic fragments only (Model 2) and MELD + peptidomic fragments (Model 3).

**Table S1 -Plasma peptides changed in AH.**

| UniProt Entry Name | Accession | Start | End | Sequence | Log2FC | -Log10PV |
|---|---|---|---|---|---|---|
| **Plasma peptides significantly changed in AH-moderate vs healthy control.** | | | | | | |
| A1AT | P01009 | 405 | 418 | SPLFMGKVVNPTQK | 1.44 | 2.49 |
| ACTB | P60709 | 99 | 105 | EEHPVLL | 1.47 | 2.91 |
| AGRG6 | Q86SQ4 | 841 | 854 | THFGVLM(+15.99)DLPRSAS | 1.19 | 1.98 |
| AMBP | P02760 | 338 | 352 | GVPGDGDEELLRFSN | 1.19 | 3.10 |
| AP1B1 | Q10567 | 664 | 676 | D(-18.01)SLIGGT(-18.01)NFVAPP | 1.90 | 3.65 |
| APOC2 | P02655 | 91 | 101 | DQVLSVLKGEE | 1.86 | 3.43 |
| APOL1 | O14791 | 28 | 51 | EEAGARVQQNVPSGTDTGDPQSKP | 1.69 | 5.27 |
| BHE22 | Q8NFJ8 | 80 | 101 | PGGGGGGSAGSGGGGGGGVGVP | 2.05 | 3.22 |
| BIN2 | Q9UBW5 | 259 | 272 | SLVISPPVRTATVS | 1.40 | 2.61 |
| | | 541 | 565 | M(+15.99)VPENNNLTAPEPQEEVSTSENPQL | 1.49 | 2.04 |
| CAVN2 | O95810 | 2 | 15 | G(+42.01)EDAAQAEKFQHPG | 5.62 | 4.96 |
| | | 350 | 360 | EIAEEAAEKAT | 2.00 | 6.64 |
| | | 395 | 425 | YEGSYALTSEEAERSDGDPVQPAVLQVHQTS | 4.17 | 5.08 |
| | | 403 | 425 | SEEAERSDGDPVQPAVLQVHQTS | 3.04 | 3.36 |
| | | 406 | 425 | AERSDGDPVQPAVLQVHQTS | 1.54 | 1.82 |
| | | 407 | 425 | ERSDGDPVQPAVLQVHQTS | 1.45 | 2.55 |
| | | 411 | 425 | GDPVQPAVLQVHQTS | 1.99 | 1.41 |
| | | 413 | 425 | PVQPAVLQVHQTS | 1.45 | 3.59 |
| CCG8 | Q8WXS5 | 337 | 371 | GGAAGGAGGGGGGGGGAGAERDRGGASGFLTLHNA | 1.12 | 1.53 |
| CD99 | P14209 | 89 | 114 | SFSDADLADGVSGGEGKGGSDGGGSH | 1.10 | 3.00 |
| CO1A1 | P02452 | 230 | 249 | GDDGEAGKPGR(+15.99)PGER(+15.99)GPP(+15.99)GP | 1.44 | 4.15 |
| | | 232 | 249 | DGEAGKPGRP(+15.99)GERGPPGP(+15.99) | 2.03 | 4.76 |
| | | 233 | 249 | GEAGKPGR(+15.99)P(+15.99)GERGPP(+15.99)GP | 1.85 | 3.97 |
| | | 235 | 249 | AGKPGRP(+15.99)GERGPPGP(+15.99) | 1.60 | 1.67 |
| | | 287 | 305 | GEP(+15.99)GSP(+15.99)GENGAPGQM(+31.99)GPRG | 2.15 | 3.97 |
| | | 340 | 368 | AGPP(+15.99)GFP(+15.99)GAVGAK(+15.99)GEAGPQGPRGSEGPQG | 1.56 | 2.89 |

| UniProt Entry Name | Accession | Start | End | Sequence | Log2FC | -Log10PV |
|---|---|---|---|---|---|---|
| | | 430 | 455 | KGNSGEP(+15.99)GAP(+15.99)GSKGDTGAK(+15.99)GEPGPVG | 2.86 | 4.67 |
| | | 431 | 455 | GNSGEP(+15.99)GAP(+15.99)GSKGDTGAKGEP(+15.99)GPVG | 3.45 | 4.76 |
| | | 432 | 453 | NSGEP(+15.99)GAP(+15.99)GSKGDTGAKGEPGP(+15.99) | 1.52 | 1.83 |
| | | 439 | 453 | P(+15.99)GSKGDTGAKGEPGP(+15.99) | 1.43 | 2.79 |
| | | 524 | 539 | GEAGRP(+15.99)GEAGLP(+15.99)GAKG | 1.45 | 3.74 |
| | | 540 | 558 | LTGSP(+15.99)GSP(+15.99)GPDGKTGPP(+15.99)GP | 1.86 | 1.74 |
| | | 543 | 558 | SP(+15.99)GSP(+15.99)GPDGKTGPP(+15.99)GP | 2.71 | 1.72 |
| | | 769 | 784 | IGPP(+15.99)GPAGAP(+15.99)GDKGES | 1.15 | 2.25 |
| | | 815 | 843 | GPPGADGQPGAKGEP(+15.99)GDAGAKGDAGPPGP | 3.43 | 9.19 |
| | | 815 | 843 | GPP(+15.99)GADGQP(+15.99)GAKGEPGDAGAKGDAGPP(+15.99)GP | -3.26 | 3.16 |
| | | 818 | 843 | GADGQPGAKGEP(+15.99)GDAGAKGDAGPP(+15.99)GP | -2.89 | 3.02 |
| | | 819 | 844 | ADGQPGAKGEP(+15.99)GDAGAKGDAGPPGP(+15.99)A | 3.23 | 3.42 |
| | | 819 | 843 | ADGQPGAKGEP(+15.99)GDAGAKGDAGPP(+15.99)GP | -2.56 | 1.61 |
| | | 819 | 843 | ADGQP(+15.99)GAKGEP(+15.99)GDAGAKGDAGPP(+15.99)GP | -2.57 | 1.52 |
| | | 819 | 839 | ADGQP(+15.99)GAKGEP(+15.99)GDAGAKGD(-18.01)AG | 2.04 | 6.90 |
| | | 820 | 843 | DGQP(+15.99)GAKGEP(+15.99)GDAGAKGDAGPP(+15.99)GP | 3.49 | 1.71 |
| | | 820 | 843 | DGQPGAKGEP(+15.99)GDAGAKGDAGPP(+15.99)GP | -3.45 | 3.43 |
| | | 889 | 909 | SGNAGPP(+15.99)GPPGP(+15.99)AGKEGGKGP | 3.95 | 8.61 |
| | | 979 | 999 | SGEPGK(+15.99)QGPSGASGERGPP(+15.99)GP | 1.57 | 2.10 |
| | | 979 | 999 | SGEPGKQGPSGASGERGPP(+15.99)GP | 1.51 | 2.32 |
| | | 1012 | 1041 | SGREGAP(+15.99)GAEGSP(+15.99)GRDGSP(+15.99)GAKGDRGETGP | 1.29 | 2.66 |
| | | 1018 | 1041 | P(+15.99)GAEGSPGRD(+15.99)GSP(+15.99)GAKGDRGETGP | 1.29 | 2.01 |
| | | 1021 | 1041 | EGSP(+15.99)GRDGSP(+15.99)GAKGDRGETGP | 1.09 | 1.93 |
| | | 1042 | 1071 | AGPP(+15.99)GAPGAP(+15.99)GAPGPVGPAGKSGDRGETGP | 3.87 | 7.77 |
| | | 1177 | 1195 | VGPP(+15.99)GPP(+15.99)GPP(+15.99)GPPGPPSAG | -1.55 | 1.50 |
| | | 1177 | 1193 | VGPP(+15.99)GPPGPP(+15.99)GPPGPPS | 1.73 | 2.05 |
| CO1A2 | P08123 | 455 | 472 | SP(+15.99)GNIGPAGKEGPVGLP(+15.99)G | 1.56 | 2.33 |
| | | 612 | 635 | SGP(+15.99)PGPDGNKGEP(+15.99)GVVGAVGTAGP | 1.88 | 1.55 |
| CO3 | P01024 | 1312 | 1319 | HWESASLL | -3.22 | 2.83 |

| UniProt Entry Name | Accession | Start | End | Sequence | Log2FC | -Log10PV |
|---|---|---|---|---|---|---|
| | | 1321 | 1337 | SEETKENEGFTVTAEGK | -2.33 | 1.53 |
| CO3A1 | P02461 | 660 | 683 | GPKGDAGAPGAPGGKGDAGAPGER | -3.74 | 4.15 |
| | | 704 | 723 | EGGKGAAGPP(+15.99)GPP(+15.99)GAAGTPG(+15.99) | 1.42 | 4.38 |
| | | 1016 | 1028 | PGSDGLPGRD(+21.98)GSP | 2.52 | 3.51 |
| CO4A | P0C0L4 | 445 | 458 | QLS(-18.01)VSAGS(-18.01)P(+15.99)HPAIA | 1.11 | 3.34 |
| | | 1159 | 1173 | EGAEPLKQRVEASIS(+21.98) | 2.03 | 3.56 |
| | | 1337 | 1349 | NGFKSHALQLNNR | 3.19 | 2.04 |
| | | 1337 | 1349 | N(+.98)GFKSHALQLNNR | 2.19 | 4.68 |
| | | 1337 | 1348 | N(+.98)GFKSHALQLNN | 2.51 | 2.23 |
| | | 1337 | 1344 | NGFKSHAL | -1.70 | 1.36 |
| | | 1338 | 1352 | G(+27.99)FKS(-18.01)HALQLNNRQIR | 3.54 | 4.97 |
| | | 1341 | 1352 | SHALQLNNRQIR | 2.39 | 1.38 |
| | | 1342 | 1352 | HALQLNNRQIR | 1.93 | 1.61 |
| | | 1343 | 1352 | ALQLNNRQIR | 2.19 | 3.03 |
| | | 1429 | 1438 | DDPDAPLQPV | 1.89 | 5.69 |
| CO4A1 | P02462 | 104 | 117 | N(+.98)PGLPGIPGQD(+21.98)GPP | 1.82 | 3.33 |
| CO4A4 | P53420 | 190 | 200 | GD(+15.99)PGLPGLPGS(+162.05) | 3.97 | 8.14 |
| CO5A2 | P05997 | 366 | 381 | GPMGPLGIPGSSGFPG | 1.37 | 1.92 |
| COCA1 | Q99715 | 2787 | 2804 | GSPGVTGPSGKPGK(+43.01)P(+15.99)GDH | 3.88 | 6.03 |
| DBNL | Q9UJU6 | 335 | 350 | EEPPEQETFYEQPPLV | 2.12 | 4.43 |
| DCTN2 | Q13561 | 183 | 210 | ATKNSKGGSGGKTTGTPPDSSLVTYELH | 1.69 | 4.24 |
| ENPL | P14625 | 14 | 25 | LTFGSVRADDEV | 1.83 | 3.63 |
| ESAM | Q96AP7 | 341 | 367 | ALPSPRLPTTDGAHPQPISPIPGGVSS | 1.09 | 2.22 |
| FETUA | P02765 | 323 | 336 | VVSLGSPSGEVSHP | 1.05 | 1.42 |
| FHOD1 | Q9Y613 | 1026 | 1058 | GEAPSNPSVPVAVSSGPGRGDADSHASM(+15.99)KSLLT | 1.43 | 4.42 |
| FIBA | P02671 | 20 | 37 | ADSGEGDFLAEGGGVRGP | 1.65 | 2.29 |
| | | 20 | 34 | ADS(+79.97)GEGDFLAEGGGV(+21.98) | -3.16 | 1.46 |
| | | 20 | 32 | ADS(+79.97)GEGDFLAEGG | 2.79 | 8.20 |
| | | 20 | 32 | ADSGEGDFLAEGG | 1.29 | 3.04 |

| UniProt Entry Name | Accession | Start | End | Sequence | Log2FC | -Log10PV |
|---|---|---|---|---|---|---|
| | | 21 | 35 | DSGEGD(-18.01)FLAEGGGVR | -2.09 | 2.16 |
| | | 21 | 34 | DS(+79.97)GEGDFLAEGGGV(+21.98) | -1.87 | 1.57 |
| | | 24 | 34 | EGDFLAEGGGV(+21.98) | 1.92 | 1.55 |
| | | 226 | 238 | M(+15.99)KPVPDLVPGNFK | 1.78 | 4.19 |
| | | 297 | 328 | SGSSGPGSTGNRNPGSSGTGGTATWKPGSSGP | 1.49 | 1.46 |
| | | 426 | 442 | REYHTEKLVTSKGDKEL | 1.94 | 3.88 |
| | | 542 | 556 | SETESRGSESGIFTN | -1.28 | 3.07 |
| | | 544 | 556 | TESRGSESGIFTN | 1.20 | 2.75 |
| | | 574 | 598 | GKSSSYSKQFTSSTSYNRGDSTFES | 3.36 | 9.33 |
| | | 576 | 602 | SSSYSKQFTSSTSYNRGDSTFESKSYK | 1.86 | 1.34 |
| | | 576 | 600 | SSSYSKQFTSSTSYNR(+.98)GDSTFESKS | -2.24 | 1.56 |
| | | 576 | 600 | SSSYSKQFTSSTSYNRGDSTFESKS | -2.57 | 1.49 |
| | | 580 | 589 | SKQFTSSTSY | 1.16 | 1.53 |
| | | 586 | 604 | STSYNRGDSTFESKSYKM(+15.99)A | 2.23 | 2.54 |
| | | 590 | 604 | NRGDSTFESKSYKMA | 2.11 | 2.43 |
| | | 592 | 604 | GDSTFESKSYKMA | 2.00 | 4.12 |
| | | 592 | 604 | GDSTFESKSYKM(+15.99)A | -2.83 | 2.19 |
| | | 600 | 628 | SYKM(+15.99)ADEAGSEADHEGTHSTKRGHAKSRP | 2.03 | 4.01 |
| | | 600 | 620 | SYKMADEAGSEADHEGTHSTK | 1.78 | 2.94 |
| | | 602 | 629 | KMADEAGSEADHEGTHSTKRGHAKSRPV | 1.03 | 1.84 |
| | | 603 | 629 | MADEAGSEADHEGTHSTKRGHAKSRPV | 2.13 | 1.37 |
| | | 603 | 628 | M(+15.99)ADEAGSEADHEGTHSTKRGHAKSRP | 2.34 | 4.73 |
| | | 603 | 624 | MADEAGSEADHEGTHSTKRGHA | 1.93 | 1.40 |
| | | 605 | 624 | DEAGSEADHEGTHSTKRGHA | -2.24 | 3.05 |
| | | 608 | 629 | GSEADHEGTHSTKRGHAKSRPV | -3.43 | 2.97 |
| | | 615 | 629 | GTHSTKRGHAKSRPV | 1.80 | 3.62 |
| FIBB | P02675 | 31 | 44 | Q(-17.03)GVNDNEEGFFSAR | 3.66 | 3.83 |
| | | 31 | 43 | Q(-17.03)GVND(-18.01)NEEGFFS(-18.01)A | 2.44 | 4.55 |
| | | 31 | 43 | Q(+.98)GVN(+.98)DNEEGFFSA | 1.24 | 2.90 |

| UniProt Entry Name | Accession | Start | End | Sequence | Log2FC | -Log10PV |
|---|---|---|---|---|---|---|
| | | 31 | 42 | Q(-17.03)GVND(-18.01)NEEGFFS(-18.01) | 2.79 | 6.48 |
| | | 31 | 42 | Q(-17.03)GVNDN(+.98)EEGFFS(-18.01) | 1.33 | 3.84 |
| | | 31 | 42 | Q(-17.03)GVNDNEE(+21.98)GFFS | -2.03 | 1.59 |
| | | 31 | 41 | Q(-17.03)GVNDNEE(+21.98)GFF | 2.86 | 4.26 |
| | | 31 | 41 | Q(-17.03)GVND(-18.01)N(+.98)EEGFF | 2.05 | 1.43 |
| | | 33 | 43 | VN(+.98)DNEEGFFSA | 4.77 | 4.77 |
| FZD8 | Q9H461 | 354 | 372 | G(+43.01)GAAAGAGAAGAGAGGPGG | 1.83 | 1.50 |
| GDIR2 | P52566 | 2 | 22 | T(+42.01)EKAPEPHVEEDDDDELDSKL | 2.83 | 3.81 |
| GTR3 | P11169 | 472 | 496 | ADRSGKDGVM(+15.99)EMNSIEPAKETTTNV | 1.42 | 5.19 |
| | | 472 | 496 | ADRSGKDGVMEMNSIEPAKETTTNV | 1.12 | 1.65 |
| | | 472 | 496 | ADRSGKDGVM(+15.99)EM(+15.99)NSIEPAKETTTNV | -1.93 | 1.40 |
| | | 475 | 496 | SGKDGVMEMNSIEPAKETTTNV | -2.04 | 1.82 |
| H15 | P16401 | 2 | 20 | S(+42.01)ETAPAETATPAPVEKSPA | -1.75 | 2.63 |
| HV315 | A0A0B4J1V0 | 20 | 34 | EVQLVESGGGLVKPG | 1.17 | 1.93 |
| IGHG1 | P01857 | 317 | 327 | NHYTQKSLSLS(-18.01) | 1.09 | 2.12 |
| INS | P01308 | 57 | 87 | EAEDLQVGQVELGGGPGAGSLQPLALEGSLQ | 1.53 | 1.91 |
| ITIH4 | Q14624 | 627 | 644 | YYLQGAKIPKPEASFSPR | 1.33 | 3.86 |
| | | 674 | 686 | GPPDVPDHAAYHP | 1.53 | 4.85 |
| K2026 | Q5HYC2 | 1637 | 1651 | VTSASASAGAQPPVL | 2.19 | 4.46 |
| KNG1 | P01042 | 381 | 389 | RPP(+15.99)GFSPFR | 2.23 | 1.99 |
| | | 381 | 387 | RPPGFSP | -2.06 | 1.88 |
| LEGL | Q3ZCW2 | 2 | 21 | A(+42.01)GSVADSDAVVKLDDGHLNN | 2.12 | 5.19 |
| LIN41 | Q2Q1W2 | 686 | 699 | FLLKFGEKGTKNGQ | 1.54 | 4.39 |
| MOES | P26038 | 468 | 495 | STPHVAEPAENEQDEQDENGAEASADLR | 1.90 | 3.55 |
| MUC16 | Q8WXI7 | 8675 | 8689 | PMATTSTLGNTSVST | 1.76 | 3.91 |
| MYH9 | P35579 | 2 | 13 | A(+42.01)QQAADKYLYVD | 1.85 | 1.94 |
| NEUG | Q92686 | 54 | 75 | KGPGPGGPGGAGVARGGAGGGP | 2.82 | 4.16 |
| PDLI1 | O00151 | 167 | 181 | TAASGVEANSRPLDH | 1.78 | 9.17 |
| | | 183 | 194 | QPPSSLVIDKES | 1.34 | 3.64 |

| UniProt Entry Name | Accession | Start | End | Sequence | Log2FC | -Log10PV |
|---|---|---|---|---|---|---|
| | | 208 | 220 | EPPKQSTSFLVLQ | 2.28 | 2.28 |
| SHOX2 | O60902 | 58 | 83 | AAGGGGGGGGGGGGGGGGGGVGGGGA | 2.58 | 1.53 |
| | | 64 | 95 | GGGGGGGGGGGGGGVGGGGAGGGAGGGRSPVR | 1.31 | 3.50 |
| SLAF5 | Q9UIB8 | 203 | 214 | KPPGTSSYEIVI | 1.38 | 2.74 |
| SRC8 | Q14247 | 76 | 101 | TGPKASHGYGGKFGVEQDRM(+15.99)DKSAVG | 1.12 | 1.38 |
| | | 303 | 312 | S(-18.01)KGFGGKYGV | -1.25 | 1.51 |
| SRGN | P10124 | 134 | 155 | RSLDRNLPSDSQDLGQHGLEED | -3.53 | 3.01 |
| | | 135 | 148 | SLDRNLPSDSQDLG | -1.67 | 2.15 |
| TBA1C | Q9BQE3 | 271 | 285 | TYAPVISAEKAYHEQ | -1.63 | 3.45 |
| | | 273 | 285 | APVISAEKAYHEQ | 1.17 | 3.81 |
| | | 358 | 373 | QPPTVVPGGDLAKVQR | 2.77 | 8.86 |
| | | 358 | 373 | Q(-17.03)PPTVVPGGDLAKVQR | -1.14 | 1.35 |
| TBB1 | Q9H4B7 | 26 | 43 | EEHGIDLAGSDRGASALQ | 2.23 | 4.91 |
| | | 428 | 451 | AKAVLEEDEEVTEEAEMEPEDKGH | 2.64 | 4.73 |
| | | 430 | 451 | AVLEEDEEVTEEAEM(+15.99)EPEDKGH | -1.87 | 2.18 |
| TBB5 | P07437 | 168 | 190 | SVVPSPKVSDTVVEPYNATLSVH | 3.18 | 6.12 |
| | | 169 | 184 | VVPSPKVSDTVVEPYN | 1.65 | 1.40 |
| | | 420 | 444 | SEYQQYQDATAEEEEDFGEEAEEEA | 1.24 | 1.76 |
| TLN1 | Q9Y490 | 434 | 448 | Q(-17.03)QYNRVGKVEHGSVA | 3.10 | 2.80 |
| | | 468 | 480 | M(+15.99)PPAQQQITSGQM(+15.99) | 1.44 | 2.96 |
| | | 468 | 478 | M(+15.99)PPAQQQITSG | 1.56 | 2.54 |
| TYB4 | P62328 | 2 | 23 | SDK(+42.01)PDM(+15.99)AEIEKFDKSKLKKTET | -1.92 | 2.24 |
| | | 2 | 16 | S(+42.01)DKPDM(+15.99)AEIEKFDKS | 3.36 | 4.46 |
| | | 2 | 12 | S(+42.01)DKPDM(+15.99)AEIEK | 3.43 | 1.46 |
| | | 3 | 23 | DKPDM(+15.99)AEIEKFDKSKLKKTET | 3.07 | 8.06 |
| | | 15 | 44 | KSKLKKTETQEKNPLPSKETIEQEKQAGES | 1.66 | 5.05 |
| | | 22 | 44 | ETQEKNPLPSKETIEQEKQAGES | 3.19 | 9.41 |
| | | 24 | 44 | QEKNPLPSKETIEQEKQAGES | 5.74 | 6.66 |
| | | 25 | 44 | EKNPLPSKETIEQEKQAGES | 2.84 | 1.39 |

| UniProt Entry Name | Accession | Start | End | Sequence | Log2FC | -Log10PV |
|---|---|---|---|---|---|---|
| | | 26 | 44 | KNPLPSKETIEQEKQAGES | 3.18 | 1.54 |
| | | 30 | 44 | PSKETIEQEKQAGES | -3.00 | 2.40 |
| VIME | P08670 | 446 | 466 | TVETRDGQVINETSQHHDDLE | 1.57 | 1.75 |
| VINC | P18206 | 859 | 899 | APPKPPLPEGEVPPPRPPPPEEKDEEFPEQKAGEVINQPM(+15.99)M(+15.99) | 1.87 | 2.71 |
| ZYX | Q15942 | 2 | 15 | A(+42.01)APRPSPAISVSVS | 3.32 | 4.26 |
| | | 36 | 56 | VNPFRPGDSEPPPAPGAQRAQ | 4.18 | 2.69 |
| | | 185 | 222 | SSTKPAAGGTAPLPPWKSPSSSQPLPQVPAPAQSQTQF | 1.71 | 1.34 |
| | | 245 | 266 | VSLANTQPRGPPASSPAPAPKF | 2.67 | 6.91 |
| | | 254 | 278 | GPPASSPAPAPKFSPVTPKFTPVAS | 1.70 | 2.33 |
| | | 280 | 319 | FSPGAPGGSGSQPNQKLGHPEALSAGTGSPQPPSFTYAQQ | 1.30 | 2.21 |
| | | 280 | 307 | FSPGAPGGSGSQPNQKLGHPEALSAGTG | 1.49 | 2.94 |
| | | 346 | 360 | GAPGPLTLKEVEELE | 3.10 | 5.94 |
| **Plasma peptides significantly changed in human severe AH vs healthy control.** | | | | | | |
| A1AT | P01009 | 25 | 48 | EDPQGDAAQKTDTSHHDQDHPTFN | -2.75 | 1.97 |
| | | 405 | 418 | SPLFMGKVVNPTQK | 1.54 | 3.25 |
| A2AP | P08697 | 29 | 39 | E(-18.01)PLGRQLTSGP | 1.11 | 1.35 |
| | | 41 | 54 | Q(-17.03)EQVSPLTLLKLGN | -1.06 | 1.96 |
| ACTB | P60709 | 96 | 113 | VAPEEHPVLLTEAPLNPK | -1.66 | 2.14 |
| | | 99 | 105 | EEHPVLL | 1.62 | 3.97 |
| AMBP | P02760 | 338 | 352 | GVPGDGDEELLRFSN | 1.61 | 6.18 |
| AP1B1 | Q10567 | 631 | 648 | DLLNLDLGPPVSGPPLAT | -1.34 | 1.88 |
| | | 664 | 676 | D(-18.01)SLIGGT(-18.01)NFVAPP | 1.90 | 4.14 |
| APOC2 | P02655 | 91 | 101 | DQVLSVLKGEE | 1.63 | 2.84 |
| APOL1 | O14791 | 28 | 51 | EEAGARVQQNVPSGTDTGDPQSKP | 2.15 | 8.93 |
| ARI1B | Q8NFD5 | 308 | 349 | GGGGGGGGGGGGSGGGGGGGGAGAGGAGAGAVAAAAAAAAAA | -2.27 | 3.96 |
| BHE22 | Q8NFJ8 | 80 | 101 | PGGGGGGSAGSGGGGGGGVGVP | 2.53 | 5.22 |
| BIN2 | Q9UBW5 | 259 | 272 | SLVISPPVRTATVS | 1.29 | 2.69 |

| UniProt Entry Name | Accession | Start | End | Sequence | Log2FC | -Log10PV |
|---|---|---|---|---|---|---|
| | | 541 | 565 | M(+15.99)VPENNNLTAPEPQEEVSTSENPQL | 1.47 | 2.26 |
| CALD1 | Q05682 | 691 | 713 | SAKPTKPAASDLPVPAEGVRNIK | -1.83 | 1.58 |
| | | 705 | 713 | PAEGVRNIK | 1.14 | 2.83 |
| | | 723 | 744 | SSPTAAGTPNKETAGLKVGVSS | -1.86 | 1.62 |
| | | 723 | 743 | SSPTAAGTPNKETAGLKVGVS | -2.55 | 3.54 |
| CAP1 | Q01518 | 273 | 286 | HVSDDM(+15.99)KTHKNPAL | 1.20 | 2.76 |
| CAVN2 | O95810 | 2 | 15 | G(+42.01)EDAAQAEKFQHPG | 5.51 | 5.14 |
| | | 178 | 195 | GAVEGKEELPDENKSLEE | -2.02 | 1.62 |
| | | 347 | 360 | VEGEIAEEAAEKAT | -2.40 | 2.60 |
| | | 350 | 360 | EIAEEAAEKAT | 2.13 | 8.01 |
| | | 395 | 425 | YEGSYALTSEEAERSDGDPVQPAVLQVHQTS | 4.75 | 6.99 |
| | | 401 | 425 | LTSEEAERSDGDPVQPAVLQVHQTS | 1.71 | 2.86 |
| | | 403 | 425 | SEEAERSDGDPVQPAVLQVHQTS | 3.68 | 5.10 |
| | | 406 | 425 | AERSDGDPVQPAVLQVHQTS | 2.12 | 3.84 |
| | | 407 | 425 | ERSDGDPVQPAVLQVHQTS | 1.75 | 3.87 |
| | | 411 | 425 | GDPVQPAVLQVHQTS | 2.20 | 2.05 |
| | | 413 | 425 | PVQPAVLQVHQTS | 1.59 | 4.32 |
| CCG8 | Q8WXS5 | 337 | 371 | GGAAGGAGGGGGGGGGAGAERDRGGASGFLTLHNA | 1.12 | 1.69 |
| CD99 | P14209 | 89 | 114 | SFSDADLADGVSGGEGKGGSDGGGSH | 1.29 | 4.67 |
| CNN2 | Q99439 | 2 | 15 | S(+42.01)STQFNKGPSYGLS | -1.23 | 1.47 |
| COL1A1 | P02452 | 229 | 249 | NGDDGEAGKPGRP(+15.99)GERGPPGP(+15.99) | -3.46 | 4.40 |
| | | 229 | 249 | N(+.98)GDDGEAGKPGRPGER(+15.99)GPPGP(+15.99) | -3.64 | 1.63 |
| | | 230 | 249 | GDDGEAGKPGR(+15.99)PGER(+15.99)GPP(+15.99)GP | 1.14 | 2.89 |
| | | 232 | 249 | DGEAGKPGRP(+15.99)GERGPPGP(+15.99) | 2.39 | 7.21 |
| | | 233 | 249 | GEAGKPGR(+15.99)P(+15.99)GERGPP(+15.99)GP | 2.55 | 7.62 |
| | | 235 | 249 | AGKPGRP(+15.99)GERGPPGP(+15.99) | 2.37 | 4.18 |
| | | 236 | 249 | GKPGRP(+15.99)GERGPPGP(+15.99) | 1.63 | 1.41 |
| | | 287 | 305 | GEP(+15.99)GSP(+15.99)GENGAPGQM(+31.99)GPRG | 2.54 | 5.91 |
| | | 340 | 368 | AGPP(+15.99)GFP(+15.99)GAVGAK(+15.99)GEAGPQGPRGSEGPQG | 2.07 | 5.60 |

| UniProt Entry Name | Accession | Start | End | Sequence | Log2FC | -Log10PV |
|---|---|---|---|---|---|---|
| | | 430 | 455 | KGNSGEP(+15.99)GAP(+15.99)GSKGDTGAK(+15.99)GEPGPVG | 3.00 | 5.37 |
| | | 431 | 455 | GNSGEP(+15.99)GAP(+15.99)GSKGDTGAKGEP(+15.99)GPVG | 3.56 | 5.00 |
| | | 431 | 453 | GNSGEP(+15.99)GAP(+15.99)GSKGDTGAKGEPGP(+15.99) | 1.05 | 3.28 |
| | | 432 | 455 | NSGEPGAP(+15.99)GSKGDTGAKGEP(+15.99)GPVG | 2.25 | 2.45 |
| | | 434 | 455 | GEP(+15.99)GAP(+15.99)GSKGDTGAKGEP(+15.99)GPVG | -1.11 | 1.73 |
| | | 439 | 453 | P(+15.99)GSKGDTGAKGEPGP(+15.99) | 1.69 | 4.40 |
| | | 488 | 504 | GGP(+15.99)GSRGFP(+15.99)GADGVAGP | 1.27 | 1.33 |
| | | 521 | 539 | GSP(+15.99)GEAGRP(+15.99)GEAGLP(+15.99)GAKG | -2.67 | 1.59 |
| | | 522 | 539 | SP(+15.99)GEAGRP(+15.99)GEAGLP(+15.99)GAKG | -2.43 | 1.55 |
| | | 523 | 539 | P(+15.99)GEAGRP(+15.99)GEAGLP(+15.99)GAKG | -1.70 | 2.04 |
| | | 524 | 539 | GEAGRP(+15.99)GEAGLP(+15.99)GAKG | 1.55 | 4.84 |
| | | 540 | 558 | LTGSP(+15.99)GSP(+15.99)GPDGKTGPP(+15.99)GP | 1.63 | 1.42 |
| | | 541 | 560 | TGSP(+15.99)GSP(+15.99)GPDGKTGPP(+15.99)GPAG | -1.33 | 1.43 |
| | | 543 | 558 | SP(+15.99)GSP(+15.99)GPDGKTGPP(+15.99)GP | 3.55 | 3.44 |
| | | 650 | 664 | GPP(+15.99)GEAGKP(+15.99)GEQGVP(+15.99) | -1.07 | 1.36 |
| | | 657 | 672 | KP(+15.99)GEQGVP(+15.99)GDLGAP(+15.99)GP | -2.43 | 3.97 |
| | | 751 | 774 | KGADGSPGKD(-18.01)GVRGLT(+79.97)GPIGPPGP(+15.99) | -1.58 | 1.49 |
| | | 769 | 784 | IGPP(+15.99)GPAGAP(+15.99)GDKGES | 1.20 | 2.62 |
| | | 815 | 843 | GPPGADGQPGAKGEP(+15.99)GDAGAKGDAGPPGP | 3.51 | 9.30 |
| | | 815 | 843 | GPP(+15.99)GADGQP(+15.99)GAKGEPGDAGAKGDAGPP(+15.99)GP | -4.48 | 6.21 |
| | | 818 | 843 | GADGQP(+15.99)GAKGEP(+15.99)GDAGAKGDAGPP(+15.99)GP | -2.86 | 3.59 |
| | | 818 | 843 | GADGQPGAKGEP(+15.99)GDAGAKGDAGPP(+15.99)GP | -4.60 | 7.97 |
| | | 819 | 846 | ADGQPGAKGEP(+15.99)GDAGAKGDAGPP(+15.99)GPAGP | 1.31 | 1.44 |
| | | 819 | 844 | ADGQPGAKGEP(+15.99)GDAGAKGDAGPPGP(+15.99)A | 4.16 | 6.12 |
| | | 819 | 844 | ADGQP(+15.99)GAKGEP(+15.99)GDAGAKGDAGPPGP(+15.99)A | -2.04 | 1.80 |
| | | 819 | 843 | ADGQ(+.98)PGAKGEP(+15.99)GDAGAKGDAGPP(+15.99)GP | 1.71 | 1.87 |
| | | 819 | 843 | ADGQP(+15.99)GAKGEP(+15.99)GDAGAKGDAGPP(+15.99)GP | -4.01 | 4.28 |
| | | 819 | 843 | ADGQPGAKGEP(+15.99)GDAGAKGDAGPP(+15.99)GP | -4.10 | 4.85 |
| | | 819 | 839 | ADGQP(+15.99)GAKGEP(+15.99)GDAGAKGD(-18.01)AG | 2.23 | 8.13 |

| UniProt Entry Name | Accession | Start | End | Sequence | Log2FC | -Log10PV |
|---|---|---|---|---|---|---|
| | | 820 | 843 | DGQP(+15.99)GAKGEP(+15.99)GDAGAKGDAGPP(+15.99)GP | 3.82 | 2.40 |
| | | 820 | 843 | DGQPGAKGEPGDAGAKGDAGPPGP(+15.99) | -3.59 | 2.47 |
| | | 820 | 843 | DGQPGAKGEP(+15.99)GDAGAKGDAGPP(+15.99)GP | -4.97 | 7.60 |
| | | 874 | 888 | TGFP(+15.99)GAAGRVGPP(+15.99)GP | -2.10 | 2.92 |
| | | 889 | 909 | SGNAGPP(+15.99)GPPGP(+15.99)AGKEGGKGP | 4.17 | 9.21 |
| | | 913 | 930 | TGPAGRP(+15.99)GEVGPP(+15.99)GPP(+15.99)GP | 1.15 | 2.80 |
| | | 917 | 930 | GRP(+15.99)GEVGPP(+15.99)GPP(+15.99)GP | -3.47 | 3.09 |
| | | 979 | 999 | SGEPGKQGPSGASGERGPP(+15.99)GP | 1.92 | 3.98 |
| | | 979 | 999 | SGEPGK(+15.99)QGPSGASGERGPP(+15.99)GP | 1.72 | 2.87 |
| | | 984 | 998 | K(+27.99)QGP(+15.99)S(-18.01)GASGERGPPG | 1.03 | 1.91 |
| | | 1007 | 1041 | GPP(+15.99)GESGREGAPGAEGSPGRDGSP(+15.99)GAKGDRGETGP | 1.35 | 2.56 |
| | | 1010 | 1041 | GESGREGAPGAEGSPGRD(+15.99)GSP(+15.99)GAKGDRGETGP | 2.06 | 2.09 |
| | | 1011 | 1042 | ESGREGAP(+15.99)GAEGSP(+15.99)GRDGSP(+15.99)GAKGDRGETGPA | -1.70 | 2.32 |
| | | 1012 | 1041 | SGREGAP(+15.99)GAEGSP(+15.99)GRDGSP(+15.99)GAKGDRGETGP | 1.22 | 2.46 |
| | | 1018 | 1041 | P(+15.99)GAEGSPGRD(+15.99)GSP(+15.99)GAKGDRGETGP | 1.58 | 3.30 |
| | | 1021 | 1041 | EGSP(+15.99)GRDGSP(+15.99)GAKGDRGETGP | 1.27 | 2.82 |
| | | 1023 | 1041 | SPGR(+15.99)DGSP(+15.99)GAKGDRGETGP | 1.09 | 2.52 |
| | | 1042 | 1071 | AGPP(+15.99)GAPGAP(+15.99)GAPGPVGPAGKSGDRGETGP | 4.27 | 9.19 |
| | | 1142 | 1161 | GPP(+15.99)GSAGAPGKDGLNGLP(+15.99)GP | -2.18 | 3.66 |
| | | 1177 | 1195 | VGPP(+15.99)GPP(+15.99)GPP(+15.99)GPPGPPSAG | -2.27 | 3.63 |
| | | 1177 | 1193 | VGPP(+15.99)GPPGPP(+15.99)GPPGPPS | 1.65 | 1.95 |
| CO1A2 | P08123 | 369 | 383 | Q(-17.03)GPP(+15.99)GPSGEEGKRGP | 1.93 | 2.28 |
| | | 375 | 397 | SGEEGKR(+.98)GPNGEAGSAGPP(+15.99)GPPG(+15.99) | -1.71 | 2.97 |
| | | 455 | 472 | SP(+15.99)GNIGPAGKEGPVGLP(+15.99)G | 1.46 | 2.28 |
| | | 612 | 635 | SGP(+15.99)PGPDGNKGEP(+15.99)GVVGAVGTAGP | 2.02 | 2.05 |
| | | 801 | 812 | SGISGPP(+15.99)GPP(+15.99)GP | 1.37 | 1.56 |
| CO3 | P01024 | 955 | 979 | EGVQKEDIPPADLSDQVPDTESETR | -2.26 | 2.03 |
| CO3A1 | P02461 | 660 | 683 | GPKGDAGAPGAPGGKGDAGAPGER | -3.56 | 3.97 |
| | | 704 | 723 | EGGKGAAGPP(+15.99)GPP(+15.99)GAAGTPG(+15.99) | 1.46 | 4.79 |

| UniProt Entry Name | Accession | Start | End | Sequence | Log2FC | -Log10PV |
|---|---|---|---|---|---|---|
| | | 1016 | 1028 | PGSDGLPGRD(+21.98)GSP | 3.22 | 5.92 |
| CO4A | P0C0L4 | 1159 | 1173 | EGAEPLKQRVEASIS(+21.98) | 2.09 | 4.18 |
| | | 1337 | 1351 | N(+.98)GFKSHALQLNNRQI | -1.48 | 2.07 |
| | | 1337 | 1349 | NGFKSHALQLNNR | 2.78 | 1.49 |
| | | 1337 | 1349 | N(+.98)GFKSHALQLNNR | 2.31 | 5.44 |
| | | 1337 | 1348 | N(+.98)GFKSHALQLNN | 2.92 | 3.30 |
| | | 1337 | 1348 | NGFKSHALQLN(+.98)N(-18.01) | 1.71 | 2.57 |
| | | 1338 | 1352 | G(+27.99)FKS(-18.01)HALQLNNRQIR | 4.08 | 7.04 |
| | | 1341 | 1352 | SHALQLNNRQIR | 3.05 | 2.65 |
| | | 1342 | 1352 | HALQLNNRQIR | 2.70 | 3.63 |
| | | 1343 | 1352 | ALQLNNRQIR | 2.24 | 3.47 |
| | | 1345 | 1351 | Q(-17.03)LNNRQI | -1.27 | 1.44 |
| | | 1429 | 1442 | DDPDAPLQPVTPLQ | -1.33 | 1.49 |
| | | 1429 | 1438 | DDPDAPLQPV | 2.05 | 6.80 |
| CO4A1 | P02462 | 104 | 117 | N(+.98)PGLPGIPGQD(+21.98)GPP | 2.07 | 4.74 |
| | | 1357 | 1372 | GEP(+15.99)GLPGPEGPPGLK(+27.99)G(+21.98) | -2.47 | 3.04 |
| CO4A4 | P53420 | 190 | 200 | GD(+15.99)PGLPGLPGS(+162.05) | 4.21 | 9.20 |
| CO5A2 | P05997 | 243 | 263 | GD(+15.99)PGPMGPIGSRGPEGPPGK(+42.01)P(-.98) | -1.45 | 1.40 |
| | | 366 | 381 | GPMGPLGIPGSSGFPG | 1.81 | 3.90 |
| | | 441 | 454 | GPPGSAGPPGSPGP | -1.30 | 1.76 |
| COCA1 | Q99715 | 2787 | 2804 | GSPGVTGPSGKPGK(+43.01)P(+15.99)GDH | 4.44 | 8.37 |
| | | 2915 | 2931 | TGSSGPRGLPGP(+15.99)PGP(+15.99)QG(+21.98) | 1.55 | 1.55 |
| CPNS1 | P04632 | 36 | 58 | A(+43.01)GGGGGGGGGGGGGGGGGGGT(-18.01)A(+21.98) | 1.43 | 1.70 |
| CRDL1 | Q9BU40 | 212 | 228 | DPPPSRQAGGLSRFPGA | -1.34 | 3.33 |
| DBNL | Q9UJU6 | 335 | 350 | EEPPEQETFYEQPPLV | 2.60 | 7.03 |
| DCTN2 | Q13561 | 183 | 210 | ATKNSKGGSGGKTTGTPPDSSLVTYELH | 1.92 | 6.10 |
| ENPL | P14625 | 14 | 25 | LTFGSVRADDEV | 1.82 | 3.83 |
| ESAM | Q96AP7 | 341 | 367 | ALPSPRLPTTDGAHPQPISPIPGGVSS | 1.05 | 2.16 |
| F13A | P00488 | 2 | 12 | S(+42.01)ETSRTAFGGR | 1.40 | 2.19 |

| UniProt Entry Name | Accession | Start | End | Sequence | Log2FC | -Log10PV |
|---|---|---|---|---|---|---|
| | | 14 | 38 | AVPPNNSNAAEDDLPTVELQGVVPR | 2.53 | 1.56 |
| | | 18 | 38 | NNSNAAEDDLPTVELQ(+.98)GVVPR | -1.48 | 2.80 |
| FETUA | P02765 | 323 | 336 | VVSLGSPSGEVSHP | 1.35 | 2.69 |
| FHOD1 | Q9Y613 | 1026 | 1058 | GEAPSNPSVPVAVSSGPGRGDADSHASM(+15.99)KSLLT | 1.53 | 5.30 |
| FIBA | P02671 | 20 | 37 | ADSGEGDFLAEGGGVRGP | 1.86 | 3.21 |
| | | 20 | 34 | ADSGEGDFLAEGGGV | 2.71 | 2.03 |
| | | 20 | 34 | ADS(+79.97)GEGDFLAEGGGV(+21.98) | -3.62 | 2.13 |
| | | 20 | 32 | ADS(+79.97)GEGDFLAEGG | 2.76 | 8.53 |
| | | 20 | 32 | ADSGEGDFLAEGG | 1.44 | 4.18 |
| | | 20 | 31 | ADSGEGDFLAEG | 1.78 | 1.47 |
| | | 21 | 35 | DSGEGDFLAEGGGVR(+.98) | 2.88 | 2.51 |
| | | 21 | 35 | DSGEGD(-18.01)FLAEGGGVR | -3.20 | 5.59 |
| | | 21 | 34 | DS(+79.97)GEGDFLAEGGGV(+21.98) | -2.38 | 3.01 |
| | | 24 | 34 | EGDFLAEGGGV(+21.98) | 2.19 | 2.34 |
| | | 25 | 34 | GDFLAEGGGV | -2.14 | 1.46 |
| | | 226 | 238 | M(+15.99)KPVPDLVPGNFK | 1.89 | 5.23 |
| | | 240 | 249 | Q(-17.03)LQKVPPEWK | -2.64 | 3.13 |
| | | 297 | 328 | SGSSGPGSTGNRNPGSSGTGGTATWKPGSSGP | 1.75 | 2.23 |
| | | 299 | 328 | SSGPGSTGNRNPGSSGTGGTATWKPGSSGP | -2.95 | 2.71 |
| | | 310 | 328 | PGSSGTGGTATWKPGSSGP | -1.74 | 2.51 |
| | | 335 | 361 | NSGSSGTGSTGNQNPGSPRPGSTGTWN | -3.06 | 2.71 |
| | | 426 | 442 | REYHTEKLVTSKGDKEL | 2.34 | 6.24 |
| | | 524 | 535 | STGKTFPGFFSP | -1.25 | 2.51 |
| | | 542 | 556 | SETESRGSESGIFTN | -1.50 | 4.69 |
| | | 544 | 556 | TESRGSESGIFTN | 1.03 | 2.16 |
| | | 574 | 598 | GKSSSYSKQFTSSTSYNRGDSTFES | 3.46 | 9.33 |
| | | 576 | 602 | SSSYSKQFTSSTSYNRGDSTFESKSYK | 1.77 | 1.37 |
| | | 582 | 604 | Q(-17.03)FTSSTSYNRGDSTFESKSYKM(+15.99)A | 1.39 | 1.61 |
| | | 582 | 603 | Q(-17.03)FTSSTSYNRGDSTFESKSYKM(+15.99) | 1.99 | 2.39 |

| UniProt Entry Name | Accession | Start | End | Sequence | Log2FC | -Log10PV |
|---|---|---|---|---|---|---|
| | | 582 | 600 | Q(-17.03)FTSSTSYNRGDSTFESKS | 1.98 | 1.40 |
| | | 582 | 599 | Q(-17.03)FTSSTSYNRGDSTFESK | 2.83 | 2.23 |
| | | 584 | 598 | TSSTSYNRGDSTFES | -1.88 | 1.41 |
| | | 586 | 604 | STSYNRGDSTFESKSYKM(+15.99)A | 2.61 | 3.78 |
| | | 586 | 603 | STSYNRGDSTFESKSYKM(+15.99) | -1.85 | 2.03 |
| | | 590 | 604 | NRGDSTFESKSYKMA | 2.47 | 3.77 |
| | | 590 | 601 | NRGDSTFESKSY | 2.41 | 2.03 |
| | | 592 | 604 | GDSTFESKSYKMA | 2.53 | 6.94 |
| | | 592 | 604 | GDSTFESKSYKM(+15.99)A | -4.35 | 5.80 |
| | | 600 | 629 | SYKM(+15.99)ADEAGSEADHEGTHSTKRGHAKSRPV | 2.71 | 1.62 |
| | | 600 | 628 | SYKM(+15.99)ADEAGSEADHEGTHSTKRGHAKSRP | 1.95 | 4.00 |
| | | 600 | 620 | SYKMADEAGSEADHEGTHSTK | 1.89 | 3.70 |
| | | 600 | 620 | SYKM(+15.99)ADEAGSEADHEGTHSTK | -1.80 | 3.58 |
| | | 602 | 629 | KMADEAGSEADHEGTHSTKRGHAKSRPV | 1.20 | 2.98 |
| | | 602 | 629 | KM(+15.99)ADEAGSEADHEGTHSTKRGHAKSRPV | -1.29 | 1.76 |
| | | 602 | 624 | KMADEAGSEADHEGTHSTKRGHA | -1.44 | 2.00 |
| | | 603 | 629 | M(+15.99)ADEAGSEADHEGTHSTKRGHAKSRPV | 3.22 | 2.30 |
| | | 603 | 629 | MADEAGSEADHEGTHSTKRGHAKSRPV | 2.72 | 2.72 |
| | | 603 | 628 | M(+15.99)ADEAGSEADHEGTHSTKRGHAKSRP | 2.70 | 6.81 |
| | | 603 | 624 | MADEAGSEADHEGTHSTKRGHA | 2.34 | 2.35 |
| | | 603 | 620 | M(+15.99)ADEAGSEADHEGTHSTK | -1.31 | 1.36 |
| | | 605 | 628 | DEAGSEADHEGTHSTKRGHAKSRP | -1.52 | 1.48 |
| | | 605 | 624 | DEAGSEADHEGTHSTKRGHA | -3.18 | 6.36 |
| | | 607 | 624 | AGSEADHEGTHSTKRGHA | -2.68 | 4.27 |
| | | 608 | 629 | GSEADHEGTHSTKRGHAKSRPV | -2.65 | 1.89 |
| | | 610 | 629 | EADHEGTHSTKRGHAKSRPV | 1.66 | 2.72 |
| | | 611 | 622 | ADHEGTHSTKRG | -1.37 | 1.65 |
| | | 615 | 629 | GTHSTKRGHAKSRPV | 1.31 | 1.97 |
| FIBB | P02675 | 31 | 44 | Q(-17.03)GVNDNEEGFFSAR | 4.32 | 5.48 |

| UniProt Entry Name | Accession | Start | End | Sequence | Log2FC | -Log10PV |
|---|---|---|---|---|---|---|
| | | 31 | 43 | Q(-17.03)GVND(-18.01)NEEGFFS(-18.01)A | 2.29 | 4.34 |
| | | 31 | 43 | Q(+.98)GVN(+.98)DNEEGFFSA | 1.10 | 2.48 |
| | | 31 | 43 | Q(-17.03)GVNDN(+.98)EEGFFS(-18.01)A | -2.85 | 1.48 |
| | | 31 | 43 | Q(-17.03)GVNDNEE(+21.98)GFFSA | -3.27 | 2.93 |
| | | 31 | 42 | Q(-17.03)GVND(-18.01)NEEGFFS(-18.01) | 3.32 | 8.74 |
| | | 31 | 42 | Q(-17.03)GVNDN(+.98)EEGFFS(-18.01) | 1.29 | 3.82 |
| | | 31 | 42 | Q(-17.03)GVNDNEE(+21.98)GFFS | -2.78 | 3.52 |
| | | 31 | 41 | Q(-17.03)GVNDNEE(+21.98)GFF | 2.95 | 5.10 |
| | | 31 | 41 | Q(-17.03)GVND(-18.01)N(+.98)EEGFF | 2.48 | 2.42 |
| | | 31 | 41 | Q(-17.03)GVNDN(+.98)EEGFF | 2.32 | 1.62 |
| | | 31 | 40 | Q(-17.03)GVNDNEEGF | 4.20 | 1.49 |
| | | 31 | 40 | Q(-17.03)GVN(+.98)DNEEGF | 2.97 | 1.60 |
| | | 33 | 43 | VN(+.98)DNEEGFFSA | 5.71 | 6.95 |
| | | 33 | 43 | VN(+.98)D(-18.01)NEEGFFSA | -1.28 | 1.40 |
| | | 33 | 43 | VNDNEEGFFSA | -2.71 | 3.38 |
| | | 34 | 43 | NDNEEGFFSA | -3.85 | 4.45 |
| FZD8 | Q9H461 | 348 | 375 | GGAGGAGGAAAGAGAAGAGAGGPGGRGE | 1.08 | 1.35 |
| | | 354 | 372 | G(+43.01)GAAAGAGAAGAGAGGPGG | 2.05 | 2.16 |
| GDIR2 | P52566 | 2 | 22 | T(+42.01)EKAPEPHVEEDDDDELDSKL | 3.42 | 6.05 |
| GPX1 | P07203 | 40 | 48 | LLIENVASL | -1.02 | 1.50 |
| GTR3 | P11169 | 472 | 496 | ADRSGKDGVM(+15.99)EMNSIEPAKETTTNV | 1.81 | 8.65 |
| | | 472 | 496 | ADRSGKDGVMEMNSIEPAKETTTNV | 1.15 | 2.00 |
| | | 472 | 496 | ADRSGKDGVM(+15.99)EM(+15.99)NSIEPAKETTTNV | -2.33 | 2.27 |
| | | 475 | 496 | SGKDGVMEMNSIEPAKETTTNV | -3.11 | 4.90 |
| H12 | P16403 | 62 | 71 | LKKALAAAGY(-.98) | -1.84 | 2.27 |
| H15 | P16401 | 2 | 20 | S(+42.01)ETAPAETATPAPVEKSPA | -1.89 | 3.32 |
| HEP2 | P05546 | 20 | 36 | GSKGPLDQLEKGGETAQ | -1.20 | 1.86 |
| HV315 | A0A0B4J1V0 | 20 | 34 | EVQLVESGGGLVKPG | 1.40 | 2.98 |
| HV373 | A0A0B4J1V6 | 20 | 35 | EVQLVESGGGLVQPGG | 1.53 | 1.89 |

| UniProt Entry Name | Accession | Start | End | Sequence | Log2FC | -Log10PV |
|---|---|---|---|---|---|---|
| IGHG1 | P01857 | 317 | 327 | NHYTQKSLSLS(-18.01) | 1.06 | 2.14 |
| INS | P01308 | 57 | 87 | EAEDLQVGQVELGGGPGAGSLQPLALEGSLQ | 1.73 | 2.86 |
| ITA2B | P08514 | 891 | 901 | Q(-17.03)IFLPEPEQPS | -2.30 | 4.02 |
| ITIH4 | Q14624 | 627 | 644 | YYLQGAKIPKPEASFSPR | 1.09 | 2.72 |
| | | 671 | 687 | GLPGPPDVPDHAAYHPF | -2.78 | 1.55 |
| | | 674 | 686 | GPPDVPDHAAYHP | 1.80 | 6.79 |
| | | 676 | 687 | PDVPDHAAYHPF | -2.33 | 2.83 |
| K2026 | Q5HYC2 | 1637 | 1651 | VTSASASAGAQPPVL | 2.34 | 5.45 |
| KNG1 | P01042 | 381 | 389 | RPP(+15.99)GFSPFR | 2.14 | 2.01 |
| | | 381 | 387 | RPPGFSP | -2.93 | 4.31 |
| KTU | Q9NVR5 | 404 | 420 | VAGAAGSGVTTLGDPEV | -1.86 | 2.56 |
| LEGL | Q3ZCW2 | 2 | 21 | A(+42.01)GSVADSDAVVKLDDGHLNN | 2.61 | 8.08 |
| LIN41 | Q2Q1W2 | 686 | 699 | FLLKFGEKGTKNGQ | 1.41 | 3.88 |
| LTBP1 | Q14766 | 512 | 523 | KEAQPGQSQVSY | 1.08 | 4.07 |
| M3K5 | Q99683 | 964 | 977 | SISLPVPVLVEDTS | -1.20 | 1.39 |
| MOES | P26038 | 468 | 495 | STPHVAEPAENEQDEQDENGAEASADLR | 2.41 | 6.10 |
| MUC16 | Q8WXI7 | 8675 | 8689 | PMATTSTLGNTSVST | 1.80 | 4.64 |
| MYADM | Q96S97 | 2 | 14 | PVTVTRTTITTTT | -2.02 | 3.12 |
| MYH9 | P35579 | 2 | 13 | A(+42.01)QQAADKYLYVD | 1.85 | 2.08 |
| | | 1936 | 1960 | RKGAGDGS(+79.97)DEEVDGKADGAEAKPAE | 1.24 | 1.33 |
| NEUG | Q92686 | 54 | 75 | KGPGPGGPGGAGVARGGAGGGP | 3.04 | 5.47 |
| NEXN | Q0ZGT2 | 228 | 241 | VM(+15.99)DDEIESEAKKES | -1.08 | 1.48 |
| PARVB | Q9HBI1 | 39 | 50 | S(+42.01)DLQEEGKNAIN | -1.35 | 1.49 |
| PDLI1 | O00151 | 152 | 164 | SSENISNFNNALE | -2.31 | 1.61 |
| | | 160 | 173 | NNALESKTAASGVE | 1.38 | 1.50 |
| | | 167 | 181 | TAASGVEANSRPLDH | 1.78 | 9.37 |
| | | 183 | 194 | QPPSSLVIDKES | 1.15 | 2.93 |
| | | 201 | 214 | Q(-17.03)EKQELNEPPKQST | 1.49 | 2.31 |
| | | 208 | 220 | EPPKQSTSFLVLQ | 2.85 | 4.26 |

| UniProt Entry Name | Accession | Start | End | Sequence | Log2FC | -Log10PV |
|---|---|---|---|---|---|---|
| | | 221 | 245 | EILESEEKGDPNKPSGFRSVKAPVT | -2.95 | 3.63 |
| PGRP2 | Q96PD5 | 564 | 574 | EPPPRTLPATD | 1.01 | 3.54 |
| PRB2 | P02812 | 361 | 416 | SPPGKPQGPPQQEGNNPQGPPPPAGGNPQQPQAPPAGQPQGPPRPPQGGRPSRPPQ | -1.26 | 2.49 |
| RTN4 | Q9NQC3 | 1 | 23 | M(+42.01)(+15.99)EDLDQSPLVSSSDS(+79.97)PPRPQPAF | 1.20 | 2.01 |
| SHOX2 | O60902 | 58 | 83 | AAGGGGGGGGGGGGGGGGGGVGGGGA | 3.35 | 3.06 |
| | | 64 | 95 | GGGGGGGGGGGGGGVGGGGAGGGAGGGRSPVR | 1.31 | 3.94 |
| SLAF5 | Q9UIB8 | 203 | 214 | KPPGTSSYEIVI | 1.46 | 3.30 |
| SRC8 | Q14247 | 76 | 101 | TGPKASHGYGGKFGVEQDRM(+15.99)DKSAVG | 1.30 | 2.16 |
| SRGN | P10124 | 134 | 155 | RSLDRNLPSDSQDLGQHGLEED | -4.77 | 5.83 |
| | | 135 | 155 | SLDRNLPSDSQDLGQHGLEED | 2.15 | 2.27 |
| | | 135 | 148 | SLDRNLPSDSQDLG | -2.02 | 3.42 |
| | | 137 | 155 | DRNLPSDSQDLGQHGLEED | -1.85 | 2.17 |
| TBA1C | Q9BQE3 | 170 | 186 | SIYPAPQVSTAVVEPYN | -1.34 | 1.34 |
| | | 271 | 285 | TYAPVISAEKAYHEQ | -1.97 | 5.59 |
| | | 273 | 285 | APVISAEKAYHEQ | 1.37 | 5.52 |
| | | 358 | 373 | QPPTVVPGGDLAKVQR | 2.44 | 8.08 |
| | | 358 | 373 | Q(-17.03)PPTVVPGGDLAKVQR | -1.20 | 1.77 |
| TBB1 | Q9H4B7 | 26 | 43 | EEHGIDLAGSDRGASALQ | 2.31 | 5.44 |
| | | 426 | 451 | Q(-17.03)DAKAVLEEDEEVTEEAEM(+15.99)EPEDKGH | 1.06 | 1.46 |
| | | 428 | 451 | AKAVLEEDEEVTEEAEMEPEDKGH | 2.58 | 4.96 |
| | | 430 | 451 | AVLEEDEEVTEEAEM(+15.99)EPEDKGH | -2.45 | 4.19 |
| TBB5 | P07437 | 168 | 190 | SVVPSPKVSDTVVEPYNATLSVH | 3.35 | 6.96 |
| | | 169 | 184 | VVPSPKVSDTVVEPYN | 1.76 | 1.82 |
| | | 420 | 444 | SEYQQYQDATAEEEEDFGEEAEEEA | 1.36 | 2.39 |
| TCOF | Q13428 | 651 | 660 | T(+43.01)(-18.01)ASAK(+27.99)VAPVR | 1.99 | 2.45 |
| TLN1 | Q9Y490 | 434 | 448 | Q(-17.03)QYNRVGKVEHGSVA | 3.22 | 3.37 |
| | | 468 | 480 | M(+15.99)PPAQQQITSGQM(+15.99) | 1.69 | 4.65 |
| | | 468 | 478 | M(+15.99)PPAQQQITSG | 1.74 | 3.55 |
| | | 2095 | 2106 | ISATK(+15.99)AAAGKVG | -1.19 | 1.56 |

| UniProt Entry Name | Accession | Start | End | Sequence | Log2FC | -Log10PV |
|---|---|---|---|---|---|---|
| | | 2512 | 2523 | ERELEEARKKLA | -1.28 | 2.38 |
| TTHY | P02766 | 135 | 147 | SYSTTAVVTNPKE | -1.80 | 3.18 |
| TYB10 | P63313 | 30 | 44 | PTKETIEQEKRSEIS | 1.63 | 2.09 |
| TYB4 | P62328 | 2 | 44 | S(+42.01)DKPDMAEIEKFDKSKLKKTETQEKNPLPSKETIEQEKQAGES | 2.38 | 1.67 |
| | | 2 | 25 | SDK(+42.01)PDMAEIEKFDKSKLKKTETQE | 2.52 | 1.50 |
| | | 2 | 23 | SDK(+42.01)PDM(+15.99)AEIEKFDKSKLKKTET | -2.86 | 5.17 |
| | | 2 | 16 | S(+42.01)DKPDM(+15.99)AEIEKFDKS | 4.03 | 6.99 |
| | | 2 | 12 | S(+42.01)DKPDM(+15.99)AEIEK | 4.24 | 2.47 |
| | | 3 | 23 | DKPDM(+15.99)AEIEKFDKSKLKKTET | 3.05 | 8.85 |
| | | 3 | 22 | DKPDMAEIEKFDKSKLKKTE | -1.37 | 1.45 |
| | | 3 | 18 | DKPDMAEIEKFDKSKL | -1.19 | 1.72 |
| | | 13 | 44 | FDKSKLKKTETQEKNPLPSKETIEQEKQAGES | -1.93 | 1.70 |
| | | 15 | 44 | KSKLKKTETQEKNPLPSKETIEQEKQAGES | 1.81 | 6.54 |
| | | 16 | 44 | SKLKKTETQEKNPLPSKETIEQEKQAGES | 2.05 | 1.73 |
| | | 22 | 44 | ETQEKNPLPSKETIEQEKQAGES | 2.93 | 9.38 |
| | | 23 | 44 | TQEKNPLPSKETIEQEKQAGES | 3.56 | 1.52 |
| | | 24 | 44 | QEKNPLPSKETIEQEKQAGES | 6.12 | 7.84 |
| | | 25 | 44 | EKNPLPSKETIEQEKQAGES | 3.46 | 2.33 |
| | | 26 | 44 | KNPLPSKETIEQEKQAGES | 4.06 | 2.86 |
| | | 26 | 41 | KNPLPSKETIEQEKQA | 1.42 | 2.60 |
| | | 30 | 44 | PSKET(-18.01)IEQEKQAGES | 1.21 | 4.30 |
| | | 30 | 44 | PSKETIEQEKQAGES | -4.08 | 4.82 |
| VIME | P08670 | 446 | 466 | TVETRDGQVINETSQHHDDLE | 1.84 | 2.64 |
| VINC | P18206 | 859 | 899 | APPKPPLPEGEVPPPRPPPPEEKDEEFPEQKAGEVINQPM(+15.99)M(+15.99) | 2.08 | 3.56 |
| | | 859 | 895 | APPKPPLPEGEVPPPRPPPPEEKDEEFPEQKAGEVIN | 2.84 | 2.55 |
| ZYX | Q15942 | 2 | 15 | A(+42.01)APRPSPAISVSVS | 3.91 | 6.45 |
| | | 36 | 56 | VNPFRPGDSEPPPAPGAQRAQ | 4.48 | 3.32 |
| | | 185 | 222 | SSTKPAAGGTAPLPPWKSPSSSQPLPQVPAPAQSQTQF | 2.02 | 2.23 |
| | | 245 | 266 | VSLANTQPRGPPASSPAPAPKF | 2.17 | 4.89 |

| UniProt Entry Name | Accession | Start | End | Sequence | Log2FC | -Log10PV |
|---|---|---|---|---|---|---|
| | | 252 | 266 | PRGPPASSPAPAPKF | 1.31 | 3.33 |
| | | 254 | 278 | GPPASSPAPAPKFSPVTPKFTPVAS | 1.47 | 1.91 |
| | | 269 | 278 | VTPKFTPVAS | -1.18 | 2.77 |
| | | 279 | 319 | KFSPGAPGGSGSQPNQKLGHPEALSAGTGSPQPPSFTYAQQ | -1.70 | 3.17 |
| | | 280 | 319 | FSPGAPGGSGSQPNQKLGHPEALSAGTGSPQPPSFTYAQQ | 1.46 | 3.17 |
| | | 280 | 316 | FSPGAPGGSGSQPNQKLGHPEALSAGTGSPQPPSFTY | 1.40 | 1.53 |
| | | 280 | 307 | FSPGAPGGSGSQPNQKLGHPEALSAGTG | 1.75 | 4.43 |
| | | 283 | 316 | GAPGGSGSQPNQKLGHPEALSAGTGSPQPPSFTY | -1.44 | 1.33 |
| | | 346 | 360 | GAPGPLTLKEVEELE | 3.56 | 8.19 |

Log2FC and -Log10PV are in comparison to values from Healthy controls.

*Some peptides mapped to homologous locations on several protein isoforms. Canonical and/or most abundant parent protein is listed.

Start and end of the peptide are based on the FASTA protein sequence associated with the accession number. Mass addition (e.g., +42.02) and mass loss (e.g., -18.01) denotes chemical modification of the amino acid, corresponding to the following:

-18.01 dehydration
-17.03 pyroglutamic acid
-0.98 amidation
+0.98 deamidation
+15.99 oxidation
+18.01 hydration
+21.98 sodiation
+27.99 formylation
+28.03 dimethylation
+42.01 acetylation
+42.02 guanidiation
+43.01 carbamylation
+79.97 phosphorylation
+156.12 4-hydroxynonenal
+162.05 hexose

**Table S2. ECM-Matrisome Annotation for Pairwise Regulated Peptides.**

| Peptidome | AH Moderate vs. Healthy Control | AH Severe vs. Healthy Control | p-value* |
|---|---|---|---|
| **Total Peptides** | 183 | 313 | |
| Non-ECM | 76 (42) | 125 (40) | 0.98[a] |
| ECM Peptides | 107 (58) | 188 (60) | |
| **Core Matrisome Peptides[a]** | 95 (89) | 164 (87) | |
| -Collagens | 54 (57) | 89 (54) | 0.88[b] |
| -ECM Glycoproteins | 39 (41) | 71 (43) | |
| -Proteoglycans | 2 (02) | 4 (03) | |
| **Matrisome-associated[a]** | 12 (11) | 24 (13) | |
| -ECM-affiliated Proteins | 5 (42) | 7 (29) | 0.81[c] |
| -ECM Regulators | 6 (50) | 15 (63) | |
| -Secreted Factors | 1 (08) | 2 (08) | |

Regulated peptides were identified using data from volcano plots of HC vs. Moderate, and HC vs Severe AH. Types of ECM protein (i.e., “matrisome”) were determined by MatrisomeAnalyzer, as described in Supplemental Methods.

*From a mixed effect logistic regression model with ECM as the dependent variable accounting for the correlation between sequences from the same peptide using a peptide-specific random intercept and adjusting for the sequence length.

[a]Percentages calculated using the number of ECM peptides as the denominator.

[b]Compares the distribution within core matrisome.

[c]Compares the distribution within Matrisome-associated.

**Table S3: GO terms for Biological Process (GO:0008150) of peptides changed in AH.**

| Term ID # | Term Description | Count | Enrichment | -Log10PV |
|---|---|---|---|---|
| **AH-Moderate vs. Healthy Controls** | | | | |
| GO:0007155 | Cell adhesion | 17 | 0.78 | 4.87 |
| GO:0007596 | Blood coagulation | 9 | 1.25 | 4.87 |
| GO:0034109 | Homotypic cell-cell adhesion | 7 | 1.61 | 4.87 |
| GO:0042060 | Wound healing | 11 | 1.05 | 4.87 |
| GO:0070527 | Platelet aggregation | 6 | 1.68 | 4.87 |
| GO:0030168 | Platelet activation | 7 | 1.4 | 4.42 |
| GO:0016043 | Cellular component organization | 36 | 0.36 | 4.38 |
| GO:0097435 | Supramolecular fiber organization | 12 | 0.86 | 3.96 |
| GO:0007010 | Cytoskeleton organization | 16 | 0.65 | 3.47 |
| GO:0098609 | Cell-cell adhesion | 11 | 0.85 | 3.32 |
| GO:0030199 | Collagen fibril organization | 5 | 1.47 | 2.92 |
| GO:0043200 | Response to amino acid | 6 | 1.25 | 2.89 |
| GO:0007160 | Cell-matrix adhesion | 6 | 1.18 | 2.57 |
| GO:0071230 | Cellular response to amino acid stimulus | 5 | 1.36 | 2.55 |
| GO:0030036 | Actin cytoskeleton organization | 10 | 0.8 | 2.54 |
| GO:0001775 | Cell activation | 11 | 0.74 | 2.49 |
| GO:0045861 | Negative regulation of proteolysis | 8 | 0.91 | 2.37 |
| GO:0006952 | Defense response | 15 | 0.57 | 2.35 |
| GO:0006950 | Response to stress | 24 | 0.39 | 2.33 |
| GO:0006953 | Acute-phase response | 4 | 1.52 | 2.30 |
| GO:0065008 | Regulation of biological quality | 25 | 0.37 | 2.28 |
| GO:0050896 | Response to stimulus | 39 | 0.24 | 2.24 |
| GO:0090066 | Regulation of anatomical structure size | 9 | 0.79 | 2.14 |
| GO:0030198 | Extracellular matrix organization | 7 | 0.94 | 2.10 |
| GO:0032501 | Multicellular organismal process | 34 | 0.26 | 1.87 |
| GO:0071417 | Cellular response to organonitrogen compound | 9 | 0.73 | 1.79 |
| GO:0007166 | Cell surface receptor signaling pathway | 17 | 0.46 | 1.76 |
| GO:0006897 | Endocytosis | 8 | 0.79 | 1.76 |
| GO:0045087 | Innate immune response | 10 | 0.66 | 1.66 |
| GO:0071495 | Cellular response to endogenous stimulus | 12 | 0.58 | 1.62 |
| GO:0002376 | Immune system process | 17 | 0.44 | 1.59 |
| GO:0051128 | Regulation of cellular component organization | 18 | 0.42 | 1.56 |
| GO:0002250 | Adaptive immune response | 7 | 0.83 | 1.55 |
| GO:0034330 | Cell junction organization | 8 | 0.75 | 1.54 |
| GO:0098542 | Defense response to other organism | 11 | 0.58 | 1.45 |
| GO:0006959 | Humoral immune response | 6 | 0.89 | 1.39 |
| GO:0022607 | Cellular component assembly | 18 | 0.4 | 1.39 |
| GO:0007015 | Actin filament organization | 6 | 0.88 | 1.36 |
| GO:0034329 | Cell junction assembly | 6 | 0.88 | 1.36 |
| GO:0035633 | Maintenance of blood-brain barrier | 3 | 1.48 | 1.34 |
| GO:0071800 | Podosome assembly | 2 | 2.14 | 1.33 |
| GO:0051248 | Negative regulation of protein metabolic process | 11 | 0.56 | 1.33 |
| GO:0032970 | Regulation of actin filament-based process | 7 | 0.78 | 1.33 |
| GO:0060627 | Regulation of vesicle-mediated transport | 8 | 0.7 | 1.30 |
| **AH-Severe vs. Healthy Controls** | | | | |
| GO:0042060 | Wound healing | 16 | 1.03 | 7.29 |
| GO:0007596 | Blood coagulation | 12 | 1.19 | 6.57 |
| GO:0007155 | Cell adhesion | 22 | 0.71 | 6.21 |
| GO:0034109 | Homotypic cell-cell adhesion | 8 | 1.47 | 5.83 |
| GO:0016043 | Cellular component organization | 51 | 0.32 | 5.34 |
| GO:0070527 | Platelet aggregation | 7 | 1.56 | 5.34 |
| GO:0030036 | Actin cytoskeleton organization | 16 | 0.82 | 5.29 |

| Term ID # | Term Description | Count | Enrichment | -Log10PV |
|---|---|---|---|---|
| GO:0007010 | Cytoskeleton organization | 23 | 0.62 | 5.29 |
| GO:0097435 | Supramolecular fiber organization | 16 | 0.80 | 5.12 |
| GO:0050878 | Regulation of body fluid levels | 13 | 0.89 | 4.80 |
| GO:0030168 | Platelet activation | 8 | 1.27 | 4.67 |
| GO:0065008 | Regulation of biological quality | 39 | 0.38 | 4.58 |
| GO:0098609 | Cell-cell adhesion | 14 | 0.76 | 3.89 |
| GO:0032970 | Regulation of actin filament-based process | 12 | 0.82 | 3.57 |
| GO:0030199 | Collagen fibril organization | 6 | 1.36 | 3.47 |
| GO:0090066 | Regulation of anatomical structure size | 13 | 0.76 | 3.46 |
| GO:0006953 | Acute-phase response | 5 | 1.43 | 2.85 |
| GO:0007160 | Cell-matrix adhesion | 7 | 1.06 | 2.68 |
| GO:0031589 | Cell-substrate adhesion | 8 | 0.96 | 2.68 |
| GO:0045861 | Negative regulation of proteolysis | 10 | 0.82 | 2.68 |
| GO:0051248 | Negative regulation of protein metabolic process | 17 | 0.56 | 2.68 |
| GO:0006950 | Response to stress | 33 | 0.34 | 2.66 |
| GO:0007015 | Actin filament organization | 9 | 0.87 | 2.60 |
| GO:0110053 | Regulation of actin filament organization | 9 | 0.86 | 2.60 |
| GO:0032956 | Regulation of actin cytoskeleton organization | 10 | 0.79 | 2.49 |
| GO:0072378 | Blood coagulation, fibrin clot formation | 4 | 1.57 | 2.48 |
| GO:0051641 | Cellular localization | 28 | 0.37 | 2.37 |
| GO:0009653 | Anatomical structure morphogenesis | 25 | 0.40 | 2.36 |
| GO:0032501 | Multicellular organismal process | 49 | 0.23 | 2.36 |
| GO:0048856 | Anatomical structure development | 42 | 0.26 | 2.36 |
| GO:0006952 | Defense response | 19 | 0.48 | 2.34 |
| GO:0051235 | Maintenance of location | 7 | 0.97 | 2.33 |
| GO:0001775 | Cell activation | 13 | 0.62 | 2.28 |
| GO:0001568 | Blood vessel development | 11 | 0.69 | 2.17 |
| GO:0043200 | Response to amino acid | 6 | 1.06 | 2.17 |
| GO:0052548 | Regulation of endopeptidase activity | 10 | 0.73 | 2.17 |
| GO:0051179 | Localization | 38 | 0.28 | 2.14 |
| GO:0051128 | Regulation of cellular component organization | 25 | 0.37 | 2.02 |
| GO:0071230 | Cellular response to amino acid stimulus | 5 | 1.17 | 2.00 |
| GO:0051493 | Regulation of cytoskeleton organization | 11 | 0.66 | 1.97 |
| GO:0050896 | Response to stimulus | 54 | 0.19 | 1.97 |
| GO:0030198 | Extracellular matrix organization | 8 | 0.81 | 1.94 |
| GO:0010770 | Positive regulation of cell morphogenesis involved in differentiation | 5 | 1.14 | 1.93 |
| GO:0002376 | Immune system process | 23 | 0.39 | 1.92 |
| GO:0072359 | Circulatory system development | 14 | 0.54 | 1.90 |
| GO:0030833 | Regulation of actin filament polymerization | 6 | 0.98 | 1.88 |
| GO:0007163 | Establishment or maintenance of cell polarity | 7 | 0.87 | 1.86 |
| GO:0051246 | Regulation of protein metabolic process | 26 | 0.35 | 1.86 |
| GO:0006996 | Organelle organization | 31 | 0.30 | 1.83 |
| GO:0010810 | Regulation of cell-substrate adhesion | 7 | 0.86 | 1.83 |
| GO:0030334 | Regulation of cell migration | 14 | 0.53 | 1.82 |
| GO:0048519 | Negative regulation of biological process | 41 | 0.24 | 1.82 |
| GO:0050793 | Regulation of developmental process | 25 | 0.35 | 1.82 |
| GO:1900026 | Positive regulation of substrate adhesion-dependent cell spreading | 4 | 1.30 | 1.82 |
| GO:0007275 | Multicellular organism development | 35 | 0.27 | 1.80 |
| GO:0030195 | Negative regulation of blood coagulation | 4 | 1.29 | 1.80 |
| GO:0034330 | Cell junction organization | 10 | 0.66 | 1.79 |
| GO:0048731 | System development | 33 | 0.28 | 1.78 |
| GO:0022604 | Regulation of cell morphogenesis | 8 | 0.76 | 1.75 |
| GO:0051651 | Maintenance of location in cell | 5 | 1.06 | 1.73 |
| GO:0045185 | Maintenance of protein location | 5 | 1.06 | 1.72 |
| GO:0007229 | Integrin-mediated signaling pathway | 5 | 1.05 | 1.70 |

| Term ID # | Term Description | Count | Enrichment | -Log10PV |
|---|---|---|---|---|
| GO:0008104 | Protein localization | 21 | 0.38 | 1.69 |
| GO:0030162 | Regulation of proteolysis | 12 | 0.56 | 1.67 |
| GO:0048514 | Blood vessel morphogenesis | 9 | 0.68 | 1.67 |
| GO:0042730 | Fibrinolysis | 3 | 1.55 | 1.66 |
| GO:1901699 | Cellular response to nitrogen compound | 11 | 0.59 | 1.63 |
| GO:0045087 | Innate immune response | 12 | 0.55 | 1.62 |
| GO:0022607 | Cellular component assembly | 24 | 0.34 | 1.57 |
| GO:0044092 | Negative regulation of molecular function | 15 | 0.47 | 1.56 |
| GO:0051017 | Actin filament bundle assembly | 4 | 1.18 | 1.54 |
| GO:0006897 | Endocytosis | 9 | 0.65 | 1.51 |
| GO:0009888 | Tissue development | 19 | 0.39 | 1.51 |
| GO:0060627 | Regulation of vesicle-mediated transport | 10 | 0.61 | 1.51 |
| GO:0045595 | Regulation of cell differentiation | 18 | 0.41 | 1.50 |
| GO:0035239 | Tube morphogenesis | 11 | 0.57 | 1.49 |
| GO:0033036 | Macromolecule localization | 23 | 0.34 | 1.48 |
| GO:0034114 | Regulation of heterotypic cell-cell adhesion | 3 | 1.45 | 1.47 |
| GO:0065007 | Biological regulation | 71 | 0.11 | 1.45 |
| GO:0033043 | Regulation of organelle organization | 15 | 0.45 | 1.43 |
| GO:0048870 | Cell motility | 14 | 0.47 | 1.42 |
| GO:0071417 | Cellular response to organonitrogen compound | 10 | 0.59 | 1.42 |
| GO:0032535 | Regulation of cellular component size | 8 | 0.68 | 1.40 |
| GO:0031532 | Actin cytoskeleton reorganization | 4 | 1.11 | 1.34 |
| GO:0007178 | Transmembrane receptor protein serine/threonine kinase signaling pathway | 6 | 0.81 | 1.30 |

Significantly changed peptides (see Figure 2) were analyzed by StringDB (see Supplemental Methods). Resulting significant GO terms were reported from that analysis..

**Table S4: GO terms for Cellular Component (GO:0005575) of peptides changed in AH.**

| Term ID # | Term Description | Count | Enrichment | -Log10PV |
|---|---|---|---|---|
| **AH-Moderate vs. Healthy Controls** | | | | |
| GO:0005788 | Endoplasmic reticulum lumen | 16 | 1.25 | 11.83 |
| GO:0005615 | Extracellular space | 35 | 0.57 | 10.46 |
| GO:0072562 | Blood microparticle | 11 | 1.51 | 10.24 |
| GO:0034774 | Secretory granule lumen | 14 | 1.18 | 9.60 |
| GO:0062023 | Collagen-containing extracellular matrix | 15 | 1.11 | 9.60 |
| GO:0005576 | Extracellular region | 37 | 0.49 | 9.40 |
| GO:0031982 | Vesicle | 36 | 0.5 | 9.34 |
| GO:0070062 | Extracellular exosome | 25 | 0.62 | 7.57 |
| GO:0098644 | Complex of collagen trimers | 6 | 1.97 | 7.57 |
| GO:0030141 | Secretory granule | 17 | 0.83 | 7.46 |
| GO:0031093 | Platelet alpha granule lumen | 7 | 1.56 | 6.74 |
| GO:0031410 | Cytoplasmic vesicle | 25 | 0.54 | 6.24 |
| GO:0012505 | Endomembrane system | 34 | 0.4 | 6.01 |
| GO:0005581 | Collagen trimer | 7 | 1.41 | 5.84 |
| GO:0071944 | Cell periphery | 38 | 0.34 | 5.74 |
| GO:0005925 | Focal adhesion | 11 | 0.96 | 5.60 |
| GO:0005912 | Adherens junction | 8 | 1.19 | 5.38 |
| GO:0005583 | Fibrillar collagen trimer | 4 | 2.06 | 5.11 |
| GO:0070161 | Anchoring junction | 17 | 0.65 | 5.06 |
| GO:0099081 | Supramolecular polymer | 15 | 0.71 | 5.06 |
| GO:0099080 | Supramolecular complex | 16 | 0.61 | 4.16 |
| GO:0005783 | Endoplasmic reticulum | 19 | 0.51 | 3.85 |
| GO:0002102 | Podosome | 4 | 1.65 | 3.74 |
| GO:0030054 | Cell junction | 19 | 0.49 | 3.60 |
| GO:0099512 | Supramolecular fiber | 13 | 0.65 | 3.59 |
| GO:0015629 | Actin cytoskeleton | 9 | 0.81 | 3.25 |
| GO:0005911 | Cell-cell junction | 9 | 0.79 | 3.14 |
| GO:0030863 | Cortical cytoskeleton | 5 | 1.23 | 3.14 |
| GO:0043229 | Intracellular organelle | 52 | 0.13 | 3.00 |
| GO:0005938 | Cell cortex | 7 | 0.9 | 2.82 |
| GO:0005737 | Cytoplasm | 49 | 0.15 | 2.72 |
| GO:0005584 | Collagen type I trimer | 2 | 2.54 | 2.68 |
| GO:0070013 | Intracellular organelle lumen | 31 | 0.28 | 2.68 |
| GO:0030864 | Cortical actin cytoskeleton | 4 | 1.28 | 2.51 |
| GO:0005856 | Cytoskeleton | 18 | 0.42 | 2.47 |
| GO:0101002 | ficolin-1-rich granule | 5 | 0.97 | 2.05 |
| GO:0005886 | Plasma membrane | 29 | 0.26 | 2.02 |
| GO:0005587 | Collagen type IV trimer | 2 | 1.99 | 1.95 |
| GO:0043227 | Membrane-bounded organelle | 50 | 0.12 | 1.94 |
| GO:0005577 | Fibrinogen complex | 2 | 1.94 | 1.87 |
| GO:1904813 | ficolin-1-rich granule lumen | 4 | 1.05 | 1.75 |
| GO:0032991 | Protein-containing complex | 28 | 0.24 | 1.71 |
| GO:0043231 | Intracellular membrane-bounded organelle | 47 | 0.13 | 1.70 |
| GO:0005622 | Intracellular anatomical structure | 53 | 0.09 | 1.66 |
| GO:0001931 | Uropod | 2 | 1.73 | 1.57 |
| GO:0001725 | Stress fiber | 3 | 1.2 | 1.50 |
| GO:0009986 | Cell surface | 9 | 0.54 | 1.50 |
| GO:0031252 | Cell leading edge | 6 | 0.69 | 1.37 |
| **AH-Severe vs. Healthy Controls** | | | | |
| GO:0005788 | Endoplasmic reticulum lumen | 21 | 1.18 | 14.82 |
| GO:0072562 | Blood microparticle | 16 | 1.48 | 14.82 |
| GO:0034774 | Secretory granule lumen | 20 | 1.14 | 13.65 |

| Term ID # | Term Description | Count | Enrichment | -Log10PV |
|---|---|---|---|---|
| GO:0005576 | Extracellular region | 55 | 0.47 | 13.55 |
| GO:0005615 | Extracellular space | 48 | 0.52 | 12.82 |
| GO:0031982 | Vesicle | 50 | 0.45 | 10.92 |
| GO:0030141 | Secretory granule | 25 | 0.81 | 10.77 |
| GO:0070062 | Extracellular exosome | 36 | 0.58 | 10.30 |
| GO:0062023 | Collagen-containing extracellular matrix | 18 | 1.00 | 10.20 |
| GO:0031091 | Platelet alpha granule | 11 | 1.44 | 10.00 |
| GO:0031093 | Platelet alpha granule lumen | 10 | 1.53 | 9.76 |
| GO:0005925 | Focal adhesion | 17 | 0.96 | 9.13 |
| GO:0070161 | Anchoring junction | 27 | 0.66 | 8.80 |
| GO:0012505 | Endomembrane system | 49 | 0.37 | 7.75 |
| GO:0015629 | Actin cytoskeleton | 16 | 0.87 | 7.32 |
| GO:0031410 | Cytoplasmic vesicle | 34 | 0.49 | 7.24 |
| GO:0030054 | Cell junction | 31 | 0.52 | 7.07 |
| GO:0098644 | Complex of collagen trimers | 6 | 1.79 | 6.78 |
| GO:0071944 | Cell periphery | 53 | 0.30 | 6.19 |
| GO:0005912 | Adherens junction | 10 | 1.10 | 6.14 |
| GO:0099081 | Supramolecular polymer | 20 | 0.65 | 5.92 |
| GO:0030863 | Cortical cytoskeleton | 8 | 1.24 | 5.70 |
| GO:0005911 | Cell-cell junction | 14 | 0.80 | 5.45 |
| GO:0030864 | Cortical actin cytoskeleton | 7 | 1.33 | 5.40 |
| GO:0005938 | Cell cortex | 11 | 0.91 | 5.06 |
| GO:0043229 | Intracellular organelle | 80 | 0.13 | 5.02 |
| GO:0043226 | Organelle | 82 | 0.12 | 4.78 |
| GO:0005581 | Collagen trimer | 7 | 1.22 | 4.72 |
| GO:0099512 | Supramolecular fiber | 18 | 0.61 | 4.64 |
| GO:0099080 | Supramolecular complex | 21 | 0.54 | 4.57 |
| GO:0005583 | Fibrillar collagen trimer | 4 | 1.87 | 4.50 |
| GO:0005737 | Cytoplasm | 75 | 0.14 | 4.43 |
| GO:0005783 | Endoplasmic reticulum | 25 | 0.44 | 4.06 |
| GO:0005856 | Cytoskeleton | 27 | 0.41 | 3.85 |
| GO:0070013 | Intracellular organelle lumen | 45 | 0.25 | 3.42 |
| GO:0005577 | Fibrinogen complex | 3 | 1.92 | 3.24 |
| GO:0005622 | Intracellular anatomical structure | 82 | 0.09 | 3.17 |
| GO:0002102 | Podosome | 4 | 1.46 | 3.16 |
| GO:0032432 | Actin filament bundle | 5 | 1.18 | 2.96 |
| GO:0005584 | Collagen type I trimer | 2 | 2.35 | 2.35 |
| GO:0031252 | Cell leading edge | 9 | 0.68 | 2.32 |
| GO:0032991 | Protein-containing complex | 41 | 0.22 | 2.24 |
| GO:0001726 | Ruffle | 6 | 0.88 | 2.19 |
| GO:0005886 | Plasma membrane | 41 | 0.22 | 2.18 |
| GO:0001725 | Stress fiber | 4 | 1.14 | 2.05 |
| GO:0043227 | Membrane-bounded organelle | 73 | 0.09 | 1.68 |
| GO:0005587 | Collagen type IV trimer | 2 | 1.81 | 1.64 |
| GO:0030016 | Myofibril | 6 | 0.75 | 1.61 |
| GO:0035578 | Azurophil granule lumen | 4 | 0.99 | 1.56 |
| GO:0043232 | Intracellular non-membrane-bounded organelle | 37 | 0.20 | 1.49 |
| GO:0140092 | bBAF complex | 2 | 1.65 | 1.44 |
| GO:0043231 | Intracellular membrane-bounded organelle | 68 | 0.10 | 1.40 |
| GO:0101002 | ficolin-1-rich granule | 5 | 0.78 | 1.35 |

Significantly changed peptides (see Figure 2) were analyzed by StringDB (see Supplemental Methods). Resulting significant GO terms were reported from that analysis.

**Table S5. Tissue enrichment terms by Brenda Tissue Ontology (BTO) of peptides changed in AH.**

| Term ID # | Term Description | Count | Enrichment | -Log10PV |
|---|---|---|---|---|
| **AH-Moderate vs. Healthy Controls** | | | | |
| BTO:0000131 | Blood plasma | 19 | 1.19 | 13.65 |
| BTO:0000132 | Blood platelet | 18 | 1.23 | 13.65 |
| BTO:0000759 | Liver | 29 | 0.67 | 10.24 |
| BTO:0000089 | Blood | 27 | 0.71 | 10.12 |
| BTO:0000570 | Hematopoietic system | 32 | 0.6 | 10.12 |
| BTO:0000345 | Digestive gland | 31 | 0.57 | 8.86 |
| BTO:0000392 | Plasma cell | 11 | 1.35 | 8.86 |
| BTO:0001486 | Skeletal system | 22 | 0.76 | 8.83 |
| BTO:0000449 | Fetus | 22 | 0.76 | 8.81 |
| BTO:0000284 | Organism form | 28 | 0.58 | 8.01 |
| BTO:0001078 | Placenta | 20 | 0.74 | 7.55 |
| BTO:0000174 | Embryonic structure | 25 | 0.56 | 6.39 |
| BTO:0003099 | Internal female genital organ | 27 | 0.52 | 6.36 |
| BTO:0001491 | Viscus | 37 | 0.38 | 6.22 |
| BTO:0000140 | Bone | 8 | 1.22 | 5.29 |
| BTO:0000141 | Bone marrow | 12 | 0.9 | 5.29 |
| BTO:0000255 | Brain cell line | 8 | 1.17 | 4.92 |
| BTO:0000988 | Pancreas | 12 | 0.82 | 4.56 |
| BTO:0000088 | Cardiovascular system | 15 | 0.69 | 4.53 |
| BTO:0000058 | Alimentary canal | 20 | 0.53 | 4.30 |
| BTO:0000421 | Connective tissue | 14 | 0.71 | 4.30 |
| BTO:0000511 | Gastrointestinal tract | 19 | 0.56 | 4.30 |
| BTO:0001539 | Parenchyma | 4 | 1.89 | 4.30 |
| BTO:0000368 | Ear | 5 | 1.45 | 3.89 |
| BTO:0001703 | Right atrium | 5 | 1.45 | 3.89 |
| BTO:0000753 | Lymphoid tissue | 17 | 0.56 | 3.80 |
| BTO:0000545 | Gut | 5 | 1.4 | 3.70 |
| BTO:0001253 | Skin | 14 | 0.62 | 3.49 |
| BTO:0000775 | Lymphocyte | 11 | 0.74 | 3.43 |
| BTO:0001279 | Spinal cord | 7 | 1.02 | 3.41 |
| BTO:0001493 | Trunk | 13 | 0.65 | 3.41 |
| BTO:0000081 | Reproductive system | 35 | 0.27 | 3.24 |
| BTO:0000580 | Blood cancer cell | 14 | 0.59 | 3.24 |
| BTO:0001271 | Leukemia cell | 13 | 0.62 | 3.24 |
| BTO:0003091 | Urogenital system | 37 | 0.26 | 3.24 |
| BTO:0000237 | Cerebrospinal fluid | 4 | 1.49 | 3.14 |
| BTO:0001489 | Whole body | 52 | 0.14 | 3.10 |
| BTO:0000634 | Integument | 18 | 0.47 | 3.00 |
| BTO:0000020 | Abdomen | 5 | 1.18 | 2.89 |
| BTO:0000083 | Female reproductive system | 33 | 0.27 | 2.89 |
| BTO:0000648 | Intestine | 14 | 0.55 | 2.89 |
| BTO:0001488 | Endocrine gland | 34 | 0.26 | 2.89 |
| BTO:0003914 | Interstitial cell of Cajal | 6 | 1.03 | 2.89 |
| BTO:0001085 | Vascular system | 8 | 0.82 | 2.85 |
| BTO:0000091 | Ascites | 4 | 1.36 | 2.77 |
| BTO:0001702 | Left atrium | 4 | 1.36 | 2.77 |
| BTO:0000562 | Heart | 10 | 0.67 | 2.66 |
| BTO:0000522 | Gland | 35 | 0.24 | 2.55 |
| BTO:0001424 | Uterus | 12 | 0.57 | 2.51 |
| BTO:0000203 | Respiratory system | 15 | 0.48 | 2.48 |
| BTO:0001368 | Thorax | 9 | 0.69 | 2.42 |
| BTO:0000439 | Eye | 10 | 0.62 | 2.26 |

| Term ID # | Term Description | Count | Enrichment | -Log10PV |
|---|---|---|---|---|
| BTO:0000763 | Lung | 13 | 0.51 | 2.23 |
| BTO:0000782 | T-lymphocyte | 6 | 0.87 | 2.19 |
| BTO:0000744 | Lymphocytic leukemia cell | 7 | 0.76 | 2.09 |
| BTO:0000917 | Needle | 2 | 1.99 | 1.97 |
| BTO:0000452 | Fibroblast | 5 | 0.89 | 1.72 |
| BTO:0000431 | Excretory gland | 12 | 0.48 | 1.71 |
| BTO:0000772 | Lymphoblast | 5 | 0.86 | 1.60 |
| BTO:0001281 | Spleen | 7 | 0.65 | 1.47 |
| BTO:0000420 | Neck | 5 | 0.82 | 1.44 |
| BTO:0000042 | Animal | 53 | 0.08 | 1.34 |
| BTO:0000453 | Fibroblast cell line | 3 | 1.14 | 1.31 |
| **AH-Severe vs. Healthy Controls** | | | | |
| BTO:0000132 | Blood platelet | 28 | 1.24 | 22.36 |
| BTO:0000131 | Blood plasma | 29 | 1.18 | 22.23 |
| BTO:0000759 | Liver | 43 | 0.66 | 15.31 |
| BTO:0000570 | Hematopoietic system | 47 | 0.58 | 14.55 |
| BTO:0000089 | Blood | 39 | 0.68 | 14.33 |
| BTO:0000345 | Digestive gland | 46 | 0.55 | 13.13 |
| BTO:0000392 | Plasma cell | 15 | 1.29 | 11.87 |
| BTO:0001486 | Skeletal system | 31 | 0.73 | 11.82 |
| BTO:0001491 | Viscus | 57 | 0.38 | 10.14 |
| BTO:0000284 | Organism form | 37 | 0.51 | 8.52 |
| BTO:0000141 | Bone marrow | 18 | 0.88 | 8.27 |
| BTO:0001279 | Spinal cord | 13 | 1.1 | 7.92 |
| BTO:0003099 | Internal female genital organ | 37 | 0.47 | 7.39 |
| BTO:0000449 | Fetus | 25 | 0.63 | 7.12 |
| BTO:0000988 | Pancreas | 17 | 0.78 | 6.35 |
| BTO:0000174 | Embryonic structure | 32 | 0.48 | 6.24 |
| BTO:0001078 | Placenta | 23 | 0.62 | 6.22 |
| BTO:0000421 | Connective tissue | 20 | 0.67 | 6.03 |
| BTO:0001271 | Leukemia cell | 21 | 0.64 | 5.99 |
| BTO:0000511 | Gastrointestinal tract | 27 | 0.52 | 5.77 |
| BTO:0001489 | Whole body | 81 | 0.14 | 5.73 |
| BTO:0000203 | Respiratory system | 26 | 0.53 | 5.71 |
| BTO:0000255 | Brain cell line | 10 | 1.08 | 5.71 |
| BTO:0000580 | Blood cancer cell | 22 | 0.6 | 5.71 |
| BTO:0000058 | Alimentary canal | 28 | 0.49 | 5.51 |
| BTO:0000763 | Lung | 23 | 0.57 | 5.48 |
| BTO:0000088 | Cardiovascular system | 20 | 0.63 | 5.47 |
| BTO:0000545 | Gut | 7 | 1.36 | 5.41 |
| BTO:0001539 | Parenchyma | 5 | 1.79 | 5.40 |
| BTO:0000140 | Bone | 9 | 1.09 | 5.20 |
| BTO:0001488 | Endocrine gland | 53 | 0.27 | 5.15 |
| BTO:0000237 | Cerebrospinal fluid | 6 | 1.47 | 5.13 |
| BTO:0000903 | Atrium | 7 | 1.3 | 5.13 |
| BTO:0001253 | Skin | 20 | 0.59 | 4.99 |
| BTO:0000522 | Gland | 55 | 0.24 | 4.76 |
| BTO:0000753 | Lymphoid tissue | 23 | 0.51 | 4.57 |
| BTO:0001424 | Uterus | 19 | 0.58 | 4.54 |
| BTO:0001702 | Left atrium | 6 | 1.35 | 4.54 |
| BTO:0000368 | Ear | 6 | 1.34 | 4.51 |
| BTO:0001703 | Right atrium | 6 | 1.34 | 4.51 |
| BTO:0000042 | Animal | 84 | 0.09 | 3.96 |
| BTO:0001493 | Trunk | 17 | 0.57 | 3.89 |
| BTO:0000775 | Lymphocyte | 14 | 0.65 | 3.82 |
| BTO:0003914 | Interstitial cell of Cajal | 8 | 0.97 | 3.82 |

| Term ID # | Term Description | Count | Enrichment | -Log10PV |
|---|---|---|---|---|
| BTO:0000634 | Integument | 25 | 0.42 | 3.72 |
| BTO:0000081 | Reproductive system | 49 | 0.23 | 3.44 |
| BTO:0001239 | Serum | 4 | 1.55 | 3.44 |
| BTO:0000452 | Fibroblast | 8 | 0.9 | 3.40 |
| BTO:0000917 | Needle | 3 | 1.98 | 3.37 |
| BTO:0000574 | Hematopoietic cell | 16 | 0.55 | 3.31 |
| BTO:0000020 | Abdomen | 6 | 1.08 | 3.20 |
| BTO:0000751 | Leukocyte | 15 | 0.56 | 3.20 |
| BTO:0001085 | Vascular system | 10 | 0.73 | 3.07 |
| BTO:0000439 | Eye | 14 | 0.57 | 3.07 |
| BTO:0003091 | Urogenital system | 51 | 0.21 | 3.04 |
| BTO:0000562 | Heart | 13 | 0.6 | 3.00 |
| BTO:0000083 | Female reproductive system | 46 | 0.23 | 2.96 |
| BTO:0001281 | Spleen | 11 | 0.66 | 2.92 |
| BTO:0001368 | Thorax | 12 | 0.62 | 2.92 |
| BTO:0005288 | CL-48 cell | 7 | 0.89 | 2.85 |
| BTO:0000782 | T-lymphocyte | 8 | 0.81 | 2.82 |
| BTO:0000493 | Gall bladder | 4 | 1.33 | 2.74 |
| BTO:0000648 | Intestine | 17 | 0.45 | 2.48 |
| BTO:0000224 | Liver cell line | 3 | 1.55 | 2.39 |
| BTO:0000744 | Lymphocytic leukemia cell | 9 | 0.68 | 2.39 |
| BTO:0000202 | Sense organ | 15 | 0.48 | 2.34 |
| BTO:0001043 | Adult | 3 | 1.5 | 2.28 |
| BTO:0000091 | Ascites | 4 | 1.17 | 2.20 |
| BTO:0000341 | Diaphragm | 2 | 2.17 | 2.20 |
| BTO:0000420 | Neck | 7 | 0.77 | 2.20 |
| BTO:0001744 | Jugular vein | 2 | 2.17 | 2.20 |
| BTO:0001965 | Carcass | 2 | 2.17 | 2.20 |
| BTO:0001158 | Rectum | 4 | 1.14 | 2.11 |
| BTO:0001762 | Neonate | 2 | 2.05 | 2.06 |
| BTO:0002354 | Talus | 2 | 2.05 | 2.06 |
| BTO:0003513 | Hepatic stellate cell line | 2 | 2.05 | 2.06 |
| BTO:0004101 | Fascicle | 2 | 2.05 | 2.06 |
| BTO:0001419 | Urine | 3 | 1.38 | 2.00 |
| BTO:0000431 | Excretory gland | 16 | 0.41 | 1.94 |
| BTO:0001313 | Style | 2 | 1.95 | 1.94 |
| BTO:0001792 | Portal vein | 2 | 1.87 | 1.83 |
| BTO:0000975 | Ovary | 9 | 0.58 | 1.76 |
| BTO:0002168 | Juvenile | 2 | 1.81 | 1.74 |
| BTO:0003651 | Spindle cell | 2 | 1.81 | 1.74 |
| BTO:0006187 | Venous blood | 2 | 1.81 | 1.74 |
| BTO:0006188 | Arterial blood | 2 | 1.81 | 1.74 |
| BTO:0000379 | Embryo | 11 | 0.48 | 1.54 |
| BTO:0001487 | Adipose tissue | 5 | 0.8 | 1.50 |
| BTO:0003092 | Urinary system | 14 | 0.4 | 1.50 |
| BTO:0001244 | Urinary tract | 14 | 0.4 | 1.47 |
| BTO:0000450 | Fiber | 2 | 1.61 | 1.45 |
| BTO:0001243 | Shoot | 4 | 0.92 | 1.44 |
| BTO:0002045 | Capillary | 2 | 1.57 | 1.40 |
| BTO:0000575 | Hepatocyte | 2 | 1.54 | 1.34 |
| BTO:0000706 | Large intestine | 10 | 0.47 | 1.34 |

Significantly changed peptides (see Figure 2) were analyzed by StringDB (see Supplemental Methods). Resulting significant BTO terms were reported from that analysis..

**Table S6 Variable frequency in Markov Blanket (Model 2).** Frequency of variable appearances in the Markov blanket of the 90-day mortality using Leave-One-Out cross-validation on the dataset with peptidomic features only (Model 2). The peptides in shaded cells are those appearing in >90% of the Markov blankets of the 90-day mortality. *Project Variable ID*: the ID associated with a particular peptide fragment..

| Parent Gene Name | Protein Accession | # of appearances in 580 cross-validation runs | Project Variable ID |
|---|---|---|---|
| **VIM** | **P08670** | **578** | **X83A** |
| **APOC1** | **P02654** | **570** | **X54A** |
| **TUBB** | **P07437** | **535** | **X79C** |
| **CALD1** | **Q05682** | **528** | **X142A** |
| BIN2 | Q9UBW5 | 443 | X231B |
| ECM1 | Q16610 | 45 | X162A |
| CAVN2 | O95810 | 44 | X28D |
| LRBA | P50851 | 42 | X123B |
| PIGR | P01833 | 19 | X45B |
| PGRP2 | Q96PD5 | 17 | X206A |
| KNG1 | P01042 | 12 | X39D |
| MOES | P26038 | 10 | X112A |
| DREB | Q16643 | 10 | X1B |
| CO1A2 | P08123 | 10 | X80G |
| QSOX1 | O00391 | 10 | X8A |
| ALBU | P02768 | 9 | X63A |
| PDLI1 | O00151 | 9 | X7A |
| H2B1M | Q99879 | 6 | X212A |
| CASS4 | Q9NQ75 | 1 | X224A |
| APOA2 | P02652 | 1 | X53A |
| APOL1 | O14791 | 1 | X9A |

**Table S7. Variable frequency in Markov Blanket (Model 3).** Frequency of variable appearances in the Markov blanket of the 90-day mortality using Leave-One-Out cross-validation on the dataset with peptidomic + clinical features (Model 3). The peptides in shaded cells are those appearing in >90% of the Markov blankets of the 90-day mortality. Out of all clinical variables, only MELD score appeared in one or more Markov blankets. *Project Variable ID*: the ID associated with a particular peptide fragment..

| Parent Gene Name or Clinical Variable | Protein Accession | # of appearances in 580 cross-validation runs | Project Variable ID |
|---|---|---|---|
| TBB5 | P07437 | 570 | X79C |
| APOC1 | P02654 | 570 | X54A |
| VIME | P08670 | 564 | X83A |
| CALD1 | Q05682 | 530 | X142A |
| MELD | n/a | 334 | MELD |
| BIN2 | Q9UBW5 | 142 | X231B |
| CAVN2 | O95810 | 34 | X28D |
| PIGR | P01833 | 23 | X45B |
| ECM1 | Q16610 | 20 | X162A |
| ALT | n/a | 18 | ALT |
| PGRP2 | Q96PD5 | 17 | X206A |
| QSOX1 | O00391 | 12 | X8A |
| CO1A2 | P08123 | 10 | X80G |
| KNG1 | P01042 | 10 | X39D |
| DREB | Q16643 | 10 | X1B |
| MOES | P26038 | 10 | X112A |
| PDLI1 | O00151 | 9 | X7A |
| H2B1M | Q99879 | 6 | X212A |
| LRBA | P50851 | 5 | X123B |
| CASS4 | Q9NQ75 | 4 | X224A |
| HEP2 | P05546 | 1 | X74A |
| APOA2 | P02652 | 1 | X53A |

**Table S8. Demographics and clinical characteristics of the predicted 90-day survival by Model 1.** *p-value*: refers to statistical differences between predicted survivors and deceased across clinical characteristics. Predictions are based on Model 1 (MELD score) (see, also, **Figure 6**). Significant *p-values* are presented in **bold**..

| **Predicted Class** | **Predicted Alive** | **Predicted Deceased** | **p-value** |
|---|---|---|---|
| N | 36 | 22 | |
| Sex = Female (%) | 14 (40.0) | 6 (27.3) | 0.49 |
| Encephalopathy (%) | | | 0.03 |
| None | 30 (88.2) | 13 (59.1) | |
| Grade 1-2 (precipitant-induced) | 4 (11.8) | 8 (36.4) | |
| Grade 3-4 (chronic) | 0 ( 0.0) | 1 ( 4.5) | |
| Race (%) | | | 0.44 |
| White | 33 (94.3) | 20 (90.9) | |
| African American/Black | 2 ( 5.7) | 1 ( 4.5) | |
| Asian/Asian American | 0 ( 0.0) | 1 ( 4.5) | |
| Ascites (%) | | | <0.01 |
| None | 15 (44.1) | 1 ( 4.5) | |
| Mild/Moderate (easily managed) | 19 (55.9) | 15 (68.2) | |
| Severe (refractory despite therapy) | 0 ( 0.0) | 6 (27.3) | |
| Flight (%) | | | 0.37 |
| 1 | 8 (22.2) | 2 ( 9.1) | |
| 2 | 9 (25.0) | 4 (18.2) | |
| 3 | 6 (16.7) | 7 (31.8) | |
| 4 | 13 (36.1) | 9 (40.9) | |
| Age (median [IQR]) | 51.0 [46.0, 57.0] | 50.0 [38.2, 56.0] | 0.42 |
| MELD (median [IQR]) | 19.0 [15.5, 20.5] | 27.0 [26.0, 29.8] | <0.01 |
| Creatinine (median [IQR]) | 0.7 [0.5, 0.8] | 0.9 [0.6, 1.6] | 0.02 |
| Total Protein (median [IQR]) | 6.1 [5.5, 6.8] | 5.9 [5.5, 6.4] | 0.57 |
| Albumin (median [IQR]) | 2.6 [2.4, 2.9] | 2.5 [2.3, 3.0] | 0.62 |
| AST (median [IQR]) | 114.0 [78.5, 170.0] | 117.5 [98.5, 187.0] | 0.35 |
| ALT (median [IQR]) | 42.0 [26.5, 71.5] | 39.0 [33.2, 60.0] | 0.91 |
| AP (median [IQR]) | 148.0 [113.5, 215.0] | 155.5 [133.5, 203.0] | 0.97 |

**Table S9. Demographics and clinical characteristics of the predicted 90-day survival by Model 2.** *p-value*: refers to statistical differences between predicted survivors and deceased across clinical characteristics. Predictions are based on Model 2 (peptidomic features) (see, also, **Figure 6**). Significant *p-values* are presented in **bold**..

| Predicted Class | Predicted Alive | Predicted Deceased | p-value |
|---|---|---|---|
| N | 32 | 26 | |
| Sex = Female (%) | 13 (41.9) | 7 (26.9) | 0.37 |
| Encephalopathy (%) | | | 0.1 |
| - None | 27 (87.1) | 16 (64.0) | |
| - Grade 1-2 (precipitant-induced) | 4 (12.9) | 8 (32.0) | |
| - Grade 3-4 (chronic) | 0 ( 0.0) | 1 ( 4.0) | |
| Race (%) | | | 0.4 |
| - White | 30 (96.8) | 23 (88.5) | |
| - African American/Black | 1 ( 3.2) | 2 ( 7.7) | |
| - Asian/Asian American | 0 ( 0.0) | 1 ( 3.8) | |
| Ascites (%) | | | 0.04 |
| - None | 12 (38.7) | 4 (16.0) | |
| - Mild/Moderate (easily managed) | 18 (58.1) | 16 (64.0) | |
| - Severe (refractory despite therapy) | 1 ( 3.2) | 5 (20.0) | |
| Flight (%) | | | 0.07 |
| - 1 | 6 (18.8) | 4 (15.4) | |
| - 2 | 9 (28.1) | 4 (15.4) | |
| - 3 | 3 ( 9.4) | 10 (38.5) | |
| - 4 | 14 (43.8) | 8 (30.8) | |
| Age (median [IQR]) | 50.0 [44.5, 58.0] | 50.0 [38.8, 55.0] | 0.48 |
| MELD (median [IQR]) | 19.0 [16.0, 23.0] | 25.5 [20.0, 27.8] | 0.01 |
| Creatinine (median [IQR]) | 0.6 [0.5, 0.9] | 0.8 [0.7, 1.2] | 0.07 |
| Total Protein (median [IQR]) | 5.9 [5.6, 6.4] | 6.1 [5.3, 6.6] | 0.68 |
| Albumin (median [IQR]) | 2.5 [2.3, 2.8] | 2.6 [2.3, 3.1] | 0.41 |
| AST (median [IQR]) | 134.0 [80.5, 188.0] | 112.5 [91.2, 158.8] | 0.97 |
| ALT (median [IQR]) | 41.0 [24.5, 72.5] | 40.5 [32.2, 64.8] | 0.87 |
| AP (median [IQR]) | 147.0 [109.0, 191.5] | 155.5 [133.5, 219.8] | 0.27 |

**Table S10. Demographics and clinical characteristics of the predicted 90-day survival by Model 3.** *p-value*: refers to statistical differences between predicted survivors and deceased across clinical characteristics. Predictions are based on Model 3 (MELD score + peptidomic features) (see, also, **Figure 6**). Significant *p-values* are presented in **bold**..

| Predicted Class | Predicted Alive | Predicted Deceased | p-value |
|---|---|---|---|
| N | 37 | 21 | |
| Sex = Female (%) | 14 (38.9) | 6 (28.6) | 0.62 |
| Encephalopathy (%) | | | 0.02 |
| - None | 31 (88.6) | 12 (57.1) | |
| - Grade 1-2 (precipitant-induced) | 4 (11.4) | 8 (38.1) | |
| - Grade 3-4 (chronic) | 0 ( 0.0) | 1 ( 4.8) | |
| Race (%) | | | 0.22 |
| - White | 35 (97.2) | 18 (85.7) | |
| - African American/Black | 1 ( 2.8) | 2 ( 9.5) | |
| - Asian/Asian American | 0 ( 0.0) | 1 ( 4.8) | |
| Ascites (%) | | | 0.01 |
| - None | 14 (40.0) | 2 ( 9.5) | |
| - Mild/Moderate (easily managed) | 20 (57.1) | 14 (66.7) | |
| - Severe (refractory despite therapy) | 1 ( 2.9) | 5 (23.8) | |
| Flight (%) | | | 0.52 |
| - 1 | 7 (18.9) | 3 (14.3) | |
| - 2 | 9 (24.3) | 4 (19.0) | |
| - 3 | 6 (16.2) | 7 (33.3) | |
| - 4 | 15 (40.5) | 7 (33.3) | |
| Age (median [IQR]) | 50.5 [44.0, 57.5] | 50.0 [41.0, 56.0] | 0.6 |
| MELD (median [IQR]) | 19.0 [15.8, 22.0] | 27.0 [25.0, 30.0] | <0.01 |
| Creatinine (median [IQR]) | 0.7 [0.5, 0.8] | 0.9 [0.7, 1.4] | 0.01 |
| Total Protein (median [IQR]) | 6.2 [5.6, 6.9] | 5.6 [5.4, 6.4] | 0.2 |
| Albumin (median [IQR]) | 2.6 [2.3, 2.9] | 2.5 [2.4, 3.0] | 0.99 |
| AST (median [IQR]) | 119.5 [79.2, 183.0] | 113.0 [95.0, 160.0] | 0.63 |
| ALT (median [IQR]) | 41.5 [26.2, 70.2] | 40.0 [33.0, 64.0] | 0.98 |
| AP (median [IQR]) | 150.5 [116.8, 193.8] | 155.0 [133.0, 224.0] | 0.5 |

**Supplemental Figure Legends.**

**Figure S1. Quality control estimation for normalized peptidomic data used for standard univariate and multivariate statistical approaches using Metaboanalyst 5.0 (https://www.metaboanalyst.ca/).** Radial distribution and box-and-whisker plots were used to evaluate the impact of data preprocessing methods on data normalization (trend toward normal Gaussian distribution). Before and after normalization radial distribution plots and bar-and-whisker plots for (left) individualized intensity-based peptidomic (m/z) values and (right) patient-specific aggregated intensity-based peptidomic (m/z) values..

**Figure S2. Distribution of the selected model variables (MELD score, 5 peptidomic features) in survivors and deceased AH patients during the training phase.** *X-axis*: 90-day survival class; *Y-axis*: MELD score value or normalized peptidomic feature value..

**Figure S3. Visualization of the distribution of selected predictive variables of 90-day survival.** In Model 1, which uses the MELD score only, the MELD variable is the only significantly different variable in the two predicted classes. In Model 2 (peptidomic features) MELD is also significant, even though it was not used in Model 2 to predict survival. The *p-value* of the MELD score is less significant in Model 2 than in the other two models where MELD was used as a predictive variable. *Red boxes* designate the variables that are used for prediction in each model..

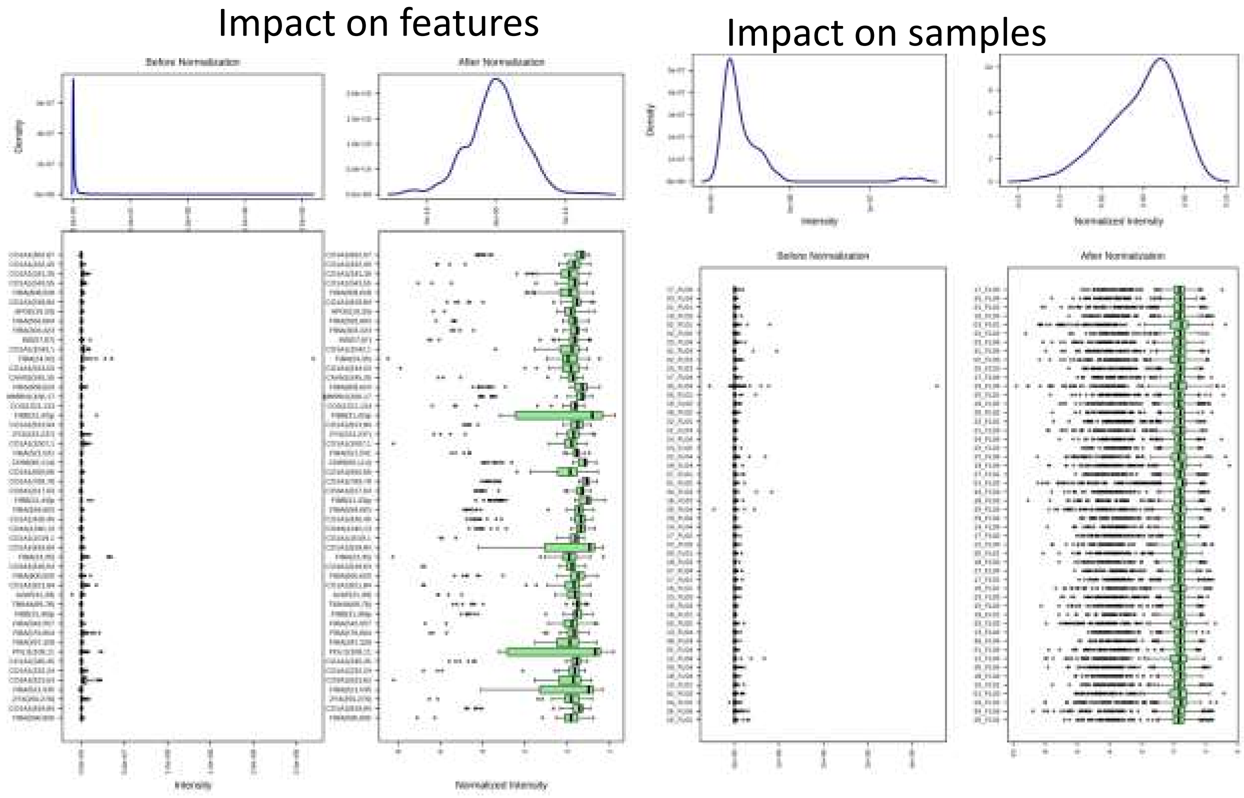


**Figure S1. Sayed et al.**

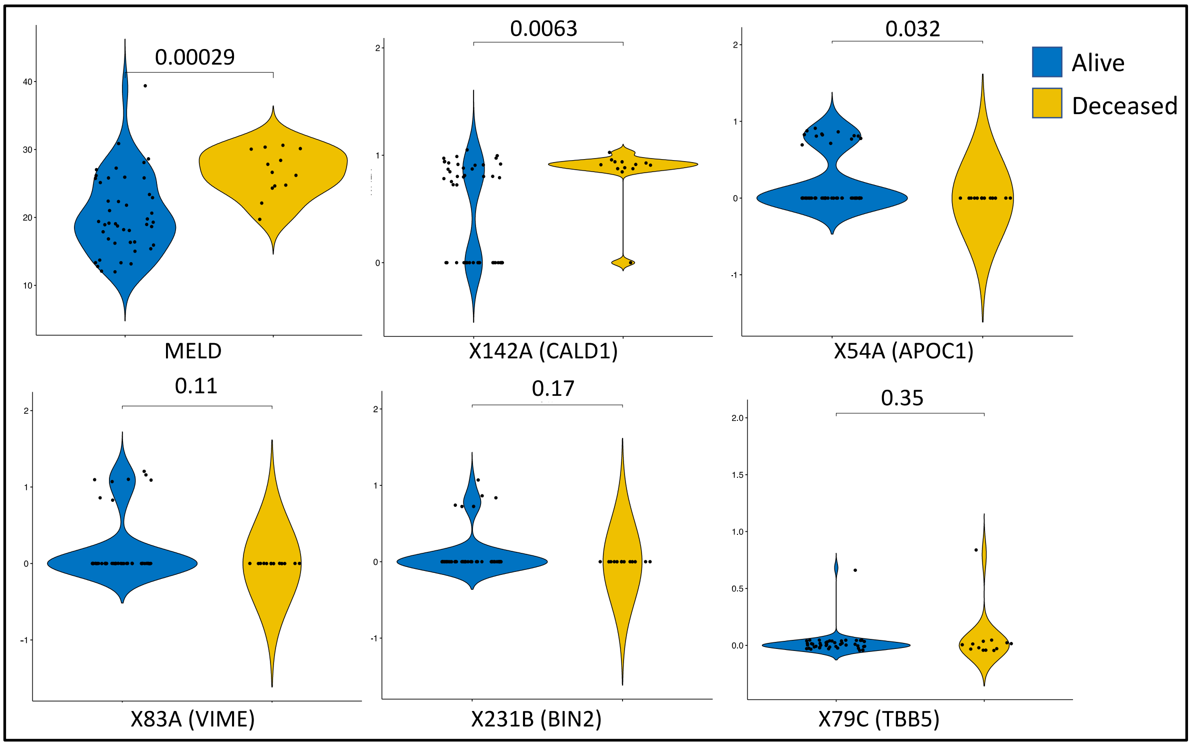


**Figure S2-Sayed et al.**

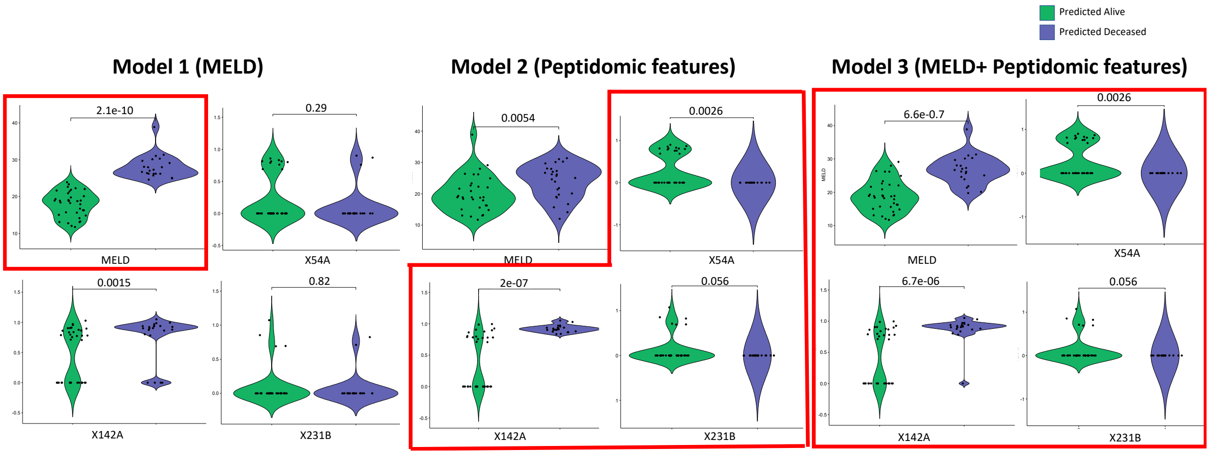


**Figure S3-Sayed et al.**
